# Prevalence of Mendelian kidney disease among patients with high-risk *APOL1* genotypes undergoing commercial genetic testing in the United States

**DOI:** 10.1101/2024.02.13.24302777

**Authors:** Ronaldo da Silva Francisco, Sumit Punj, Lisa Vincent, Nina Sanapareddy, Vivek Bhalla, Glenn M. Chertow, Dianne Keen-Kim, Vivek Charu

**Affiliations:** Department of Pathology, Stanford University School of Medicine, Stanford, CA 943051.; Natera, Inc. 201 Industrial Boulevard, San Carlos CA 94070; Division of Nephrology, Department of Medicine, Stanford University School of Medicine, Stanford, CA 94305

## Abstract

**Background:** Among individuals with high-risk *APOL1* genotypes, the lifetime risk of developing kidney failure is ∼15%, indicating that other genetic variants or non-genetic modifiers likely contribute substantially to an individual patient’s risk of progressive kidney disease. Here we estimate the prevalence and distribution of molecularly diagnosed Mendelian kidney diseases among patients with high-risk *APOL1* genotypes undergoing commercial genetic testing in the United States.

**Methods:** We analyzed clinical exome sequencing data from 15,181 individuals undergoing commercial genetic testing for Mendelian kidney disease in the United States from 2020-2021. We identified patients with high-risk *APOL1* genotypes by the presence of G1/G1, G1/G2, or G2/G2 alleles. Patients carrying single risk *APOL1* alleles were identified as G1/G0, G2/G0; the remainder of patients were G0/G0. We estimated the prevalence and distribution of molecularly diagnosed Mendelian kidney disease stratified by *APOL1* genotype.

**Results:** Of 15,181 patients, 3119 had genetic testing results consistent with a molecular diagnosis of Mendelian kidney disease (20.5%). 1035 (6.8%) had high-risk *APOL1* genotypes. The prevalence of molecularly diagnosed Mendelian kidney diseases was lower in individuals with high-risk *APOL1* genotypes (9.2%; n=95/1035) compared to single risk *APOL1* allele carriers (14.4%; n=243/1687) and those with G0/G0 *APOL1* genotypes (22.3%; n=2781/12459). The distribution of molecularly diagnosed Mendelian kidney diseases was broadly similar among patients with and without high-risk *APOL1* genotypes.

**Conclusions:** Among patients undergoing clinical genetic testing, we found a relatively high rate of molecularly diagnosed Mendelian kidney disease in patients with high-risk *APOL1* genotypes. Mendelian kidney disease may contribute to wide variation in rates of progression observed among patients with high-risk *APOL1* genotypes.

## INTRODUCTION

Americans with recent African ancestry are at increased risk of kidney failure compared to Americans of European descent, and much of this excess risk has been attributed to *APOL1*-mediated kidney disease (1). Two risk alleles in *APOL1*, the gene encoding apolipoprotein-L1, have been identified as risk factors for the development and progression of chronic kidney disease (CKD) (2). These coding variants, termed G1 and G2, are present relatively at high frequencies among people of recent African descent in part due to the positive natural selection of these alleles as they confer a protective advantage against the trypanosomes that cause African sleeping sickness (3,4). Approximately 13% of the Americans with recent African ancestry carry high-risk *APOL1* genotypes, defined as having two high-risk alleles in *APOL1* (G1/G1, G1/G2, or G2/G2) (4). The risk of developing kidney disease is 3–30 fold higher among persons with *APOL1* high-risk genotypes compared to those not carrying any *APOL1* risk alleles (5). However, among persons with high-risk *APOL1* genotypes, the lifetime risk of kidney failure is ∼15%, indicating that other genetic variants, or non-genetic modifiers, including environmental factors, likely contribute substantially to an individual patient’s risk of kidney disease (1,2).

Recent studies have demonstrated that ∼10% of patients with CKD have Mendelian forms of kidney disease (6), and as such, clinical genetic testing to identify monogenic kidney diseases has become increasingly common (7). The interaction between *APOL1* risk alleles and other causes of kidney disease is an area of active research. For example, several studies have demonstrated that the presence of *APOL1* high-risk genotypes is associated with worse disease trajectories than for patients with otherwise unrelated forms of kidney disease, and especially glomerular diseases (e.g. membranous nephropathy, systemic lupus erythematosus) (8–10). The interaction between *APOL1* risk alleles and Mendelian kidney diseases, in contrast, has been understudied.

Here we analyze real-world data from a large, ethnically diverse cohort of over 15,000 patients undergoing commercial clinical genetic testing for Mendelian kidney disease in the United States. We characterize the estimated prevalence and distribution of molecularly diagnosed Mendelian kidney diseases in this cohort, stratified by *APOL1* genotype, with the hypothesis that Mendelian kidney diseases may contribute to kidney disease incidence and progression among persons with high-risk *APOL1* genotypes.

## METHODS

This is a cross-sectional study of consented patients undergoing commercial genetic testing from April 1, 2020 to December 30, 2021. Patients were referred for genetic testing by their respective clinical providers. Demographic and clinical information collected at the time of testing includes age, race/ethnicity (either self-reported or designated by the clinical provider), sex, transplant status (yes/no), and International Classification of Diseases (ICD) diagnostic codes. We received an exemption from institutional review board review (study ID 20099-03) from Ethical & Independent Review Services, Corte Madera, California. All data were de-identified to protect patient privacy. We excluded data from patients with missing information on age and/or sex. We also excluded persons undergoing clinical genetic testing in the setting of kidney donation.

### Designated or self-reported race/ethnicity

The following race and ethnicity categories were present on the intake form are: African American, Ashkenazi Jewish, Caucasian, East Asian, French Canadian/Cajun, Hispanic, Mediterranean, Other, Sephardic Jewish, South Asian and South-East Asian. To achieve a sufficiently large sample size for further analysis, certain race/ethnic categories, namely French Canadian/Cajun, Sephardic Jewish, and Ashkenazi Jewish, were combined with the category labeled as ‘Other’ due to the limited number of individuals.

### Sequencing

Genomic DNA was extracted from either the individual’s whole blood or saliva and processed for hybrid capture-based next generation sequencing. Massively parallel sequencing was performed at 150 base-pairs, paired-end reads on a clinical exome backbone. The percentage of coverage was determined at a minimum of 20X per gene on the panel and Sanger sequencing was used to fill in regions of low coverage. The sequencing data was aligned to the GRCh37/hg19 genome assembly, and the variants were called and annotated using a bioinformatics pipeline that applied the GATK framework (11). Single nucleotide variants (SNV), insertions and deletions (indels) as well as copy number variants (CNV) were detected by this assay. Orthogonal methods for confirmation of variants included Sanger sequencing for SNV and indels and quantitative Polymerase Chain Reaction or Multiplex-ligation dependent probe amplification for CNV.

### Genetic Ancestry Determination

The population structure and admixture analyses were conducted using the continental ancestry groups from the combined, harmonized databases of the Human Genome Diversity Project (HGDP) and the 1000 Genomes Project (1KG) (12). We included SNVs with a minor allele frequency (MAF) >0.05 and missing in fewer than 5% of individuals within our dataset to determine ancestry. We retained samples for further analysis with an overall genotype rate greater than 90% (i.e., missing fewer than 10% of selected SNVs). We performed principal component analysis (PCA) using SNPWeights version 2 (10) and PLINK v1.9 (13), and estimated global genomic ancestry proportions using Admixture v1.3 (14) focusing on the following populations: African, admixed American, Central South Asia, East Asian, and European.

In additional analyses, we assigned a single ancestry group to each individual patient. Ancestry groups were determined using a clustering method derived from the PCA dimensions. We utilized the first 10 principal components (PCs) and applied the Uniform Manifold Approximation and Projection (UMAP) algorithm, via the umap package in R (15). The UMAP outputs were clustered using the Hierarchical Density-Based Spatial Clustering of Applications with Noise (HDBSCAN) algorithm, via the dbscan R package (16). Five distinct clusters were identified, corresponding to inferred continental ancestries. These clusters aligned well with the predominant admixture proportions for their respective ancestry groups (e.g., samples clustered in the African ancestry group had majority of global genomic proportions of African genetic population; **Supplemental Figure 1**). We categorized samples with unknown designated/self-reported race/ethnicity into one of the five distinct ancestry clusters.

### Interpretation of Genetic Variants

Variants were analyzed in 343 genes associated with Mendelian forms of kidney disease (see **Supplemental Table 1**), including the risk alleles associated with *APOL1*. Detected germline variants were classified using a 5-tier classification system (B - benign, LB - likely benign, VUS - variant of uncertain significance, LP - likely pathogenic, and P - pathogenic) in accordance with the American College of Medical Genetics and Genomics and the Association for Molecular Pathology (ACMG/AMP) guidelines at the time of testing (17). As part of routine clinical care, reports of identified variants and their relevant ACMG/AMP classifications were returned to the referring clinicians and patients at the time of testing.

### Identifying patients with molecularly diagnosed Mendelian kidney diseases

Accurate diagnosis of Mendelian forms of kidney disease requires genotype-phenotype correlation based on variant classification information combined with the mode of inheritance for the disorder. We classified individuals as having molecularly diagnosed Mendelian kidney disease based on the presence of P/LP variants, meeting criteria informed by empirical data (17). We considered a test “positive” or consistent with a molecular diagnosis of Mendelian kidney disease if: (1) the individual has a heterozygous or hemizygous P/LP variant in a gene associated with a dominant inheritance pattern; or (2) the patient has two P/LP variants (compound heterozygous or homozygous) in a gene with a recessive inheritance pattern. For genes with both dominant and recessive patterns of inheritance, we considered each variant’s specific phenotype and the associated pattern of inheritance was considered. For *HBB*, only results consistent with autosomal recessive sickle cell anemia were included as “positive.” We considered as carriers those individuals harboring heterozygous P/LP variants in genes associated with an autosomal recessive (AR) inheritance were considered carriers. The remaining samples were considered negative. In general, a variant classified as pathogenic (P) or likely pathogenic (LP) have met criteria informed by empirical data such that healthcare providers can use the results of molecular testing in clinical decision making (17).

### APOL1 genotype status definition

We assessed two *APOL1* high-risk alleles: (1) G1 (NM_003661.4:c.[1024A>G;1152T>G]) defined by the presence of two missense variants p.S342G (rs73885319) and p.I384M (rs60910145) that are nearly always in linkage disequilibrium; (2) G2 (NM_003661.4:c.1164_1169del) defined by the presence of an in-frame deletion of two amino acid residues at codons 388 and 389 respectively (rs71785313). We defined *APOL1* high-risk genotypes by the presence of two high-risk alleles (G1/G1, G1/G2 or G2/G2). We defined single risk *APOL1* allele carriers by the presence of only one risk allele (G1/G0 or G2/G0). The lack of *APOL1* risk alleles was denoted as (G0/G0). We deliberately considered single risk *APOL1* allele carriers because, by definition, these patients have recent African ancestry, in contrast to patients with G0/G0 *APOL1* genotypes.

### Statistical analysis

Continuous variables were summarized by means, medians and interquartile ranges, where appropriate. Comparisons of continuous variables across groups were made via Wilcoxon’s rank sum test. We described categorical variables using proportions and compared groups using Fisher’s exact test. P-values <0.05 were considered statistically significant. Analyses were conducted in R version 4.1.2.

The primary goal of our analysis was to explore the relationship between *APOL1* genotypes (exposure) and Mendelian kidney disease (outcome). However, by definition, patients with *APOL1* risk alleles have recent African ancestry, and African ancestry may confound the relationship between *APOL1* genotype and Mendelian kidney disease. As such, we conducted a matched case-control study, matching patient with high-risk *APOL1* genotypes to patients who were either single-risk *APOL1* allele carriers or G0/G0 in a 1:1 ratio, matching on age, sex, genetic ancestry proportions from the five continental populations in our study, and the first four principal components from PCA. Matching was performed via the nearest neighbor method and generalized linear models for distance estimation using the MatchIt package in R (18). We compared the prevalence of molecularly diagnosed Mendelian kidney in the matched cohorts.

## RESULTS

15,532 persons underwent clinical genetic testing during 2020-2021, of whom 15,181 (97.7%) had sufficient data for further analysis (**Figure 1**). The median age at the time of testing was 47 years (25%, 75% range 30–62 years), with ∼50% of the patients identified as female (**Table 1**). Fewer than 5% (n=696) had received a kidney transplant prior to the time of testing. Based on the designated race/ethnicity of each patient, the majority of samples were collected from Caucasian (37.1%, n=5,718), followed by 19.2% African American (n=2,960), and 12.9% Hispanic (n=1,989) persons, while the remaining (29.7%, n=4514) samples were collected from persons from other race/ethnicity categories (**Table 1**). 241 (1.6%) individuals had a multiracial/multiethnic designation. We characterized genomic ancestry using a panel of ancestry informative markers from the clinical exome sequencing data. Overall, self-designated race/ethnicity was highly concordant with genetic ancestry (**Supplemental Figure 1 and Supplemental Tables 2-4**).

**Figure 1.**
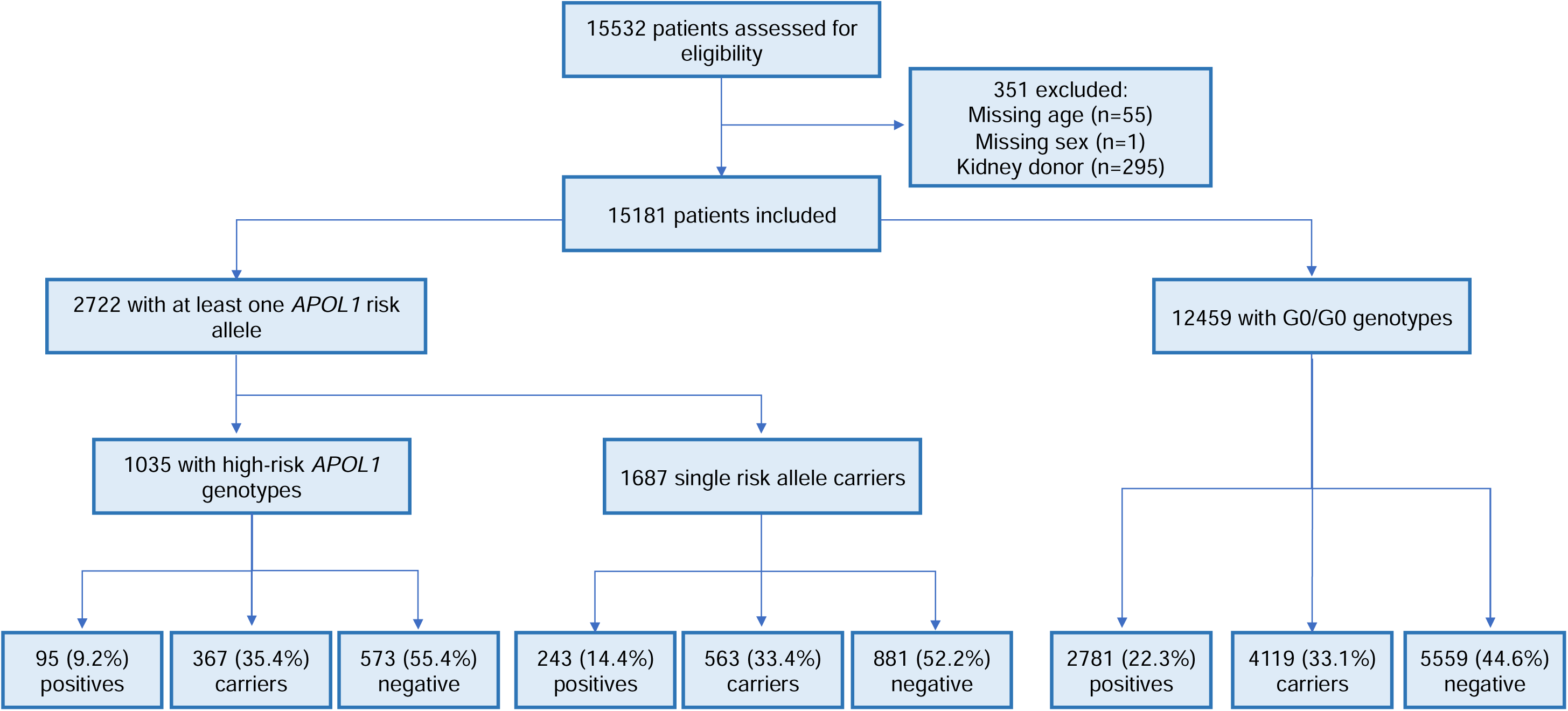
Flowchart of patients included in this study. Positive samples refer to the number of patients molecularly diagnosed with Mendelian kidney disease. Carriers were defined as patients harboring heterozygous P/LP (pathogenic/likely pathogenic) variants in genes associated with a recessive inheritance pattern. The remaining samples were considered negative.

**Figure 2.**
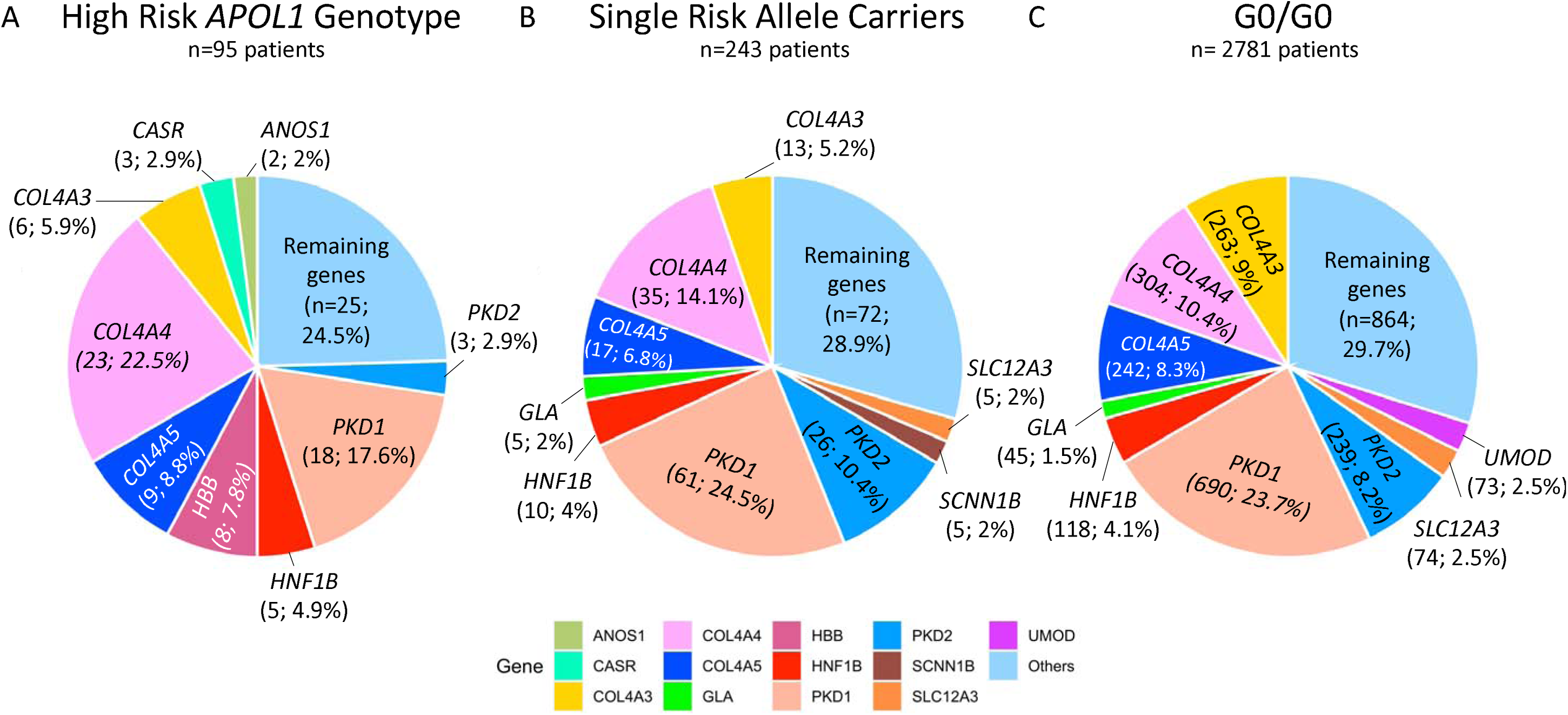
Most common genes among patients with molecularly diagnosed Mendelian kidney diseases stratified by *APOL1* genotype. Proportional representation (n, %) of most common genes in patients with (a) high-risk APOL1 genotypes, (b) single-risk allele carriers and (c) G0/G0 patients.

**Table 1.**
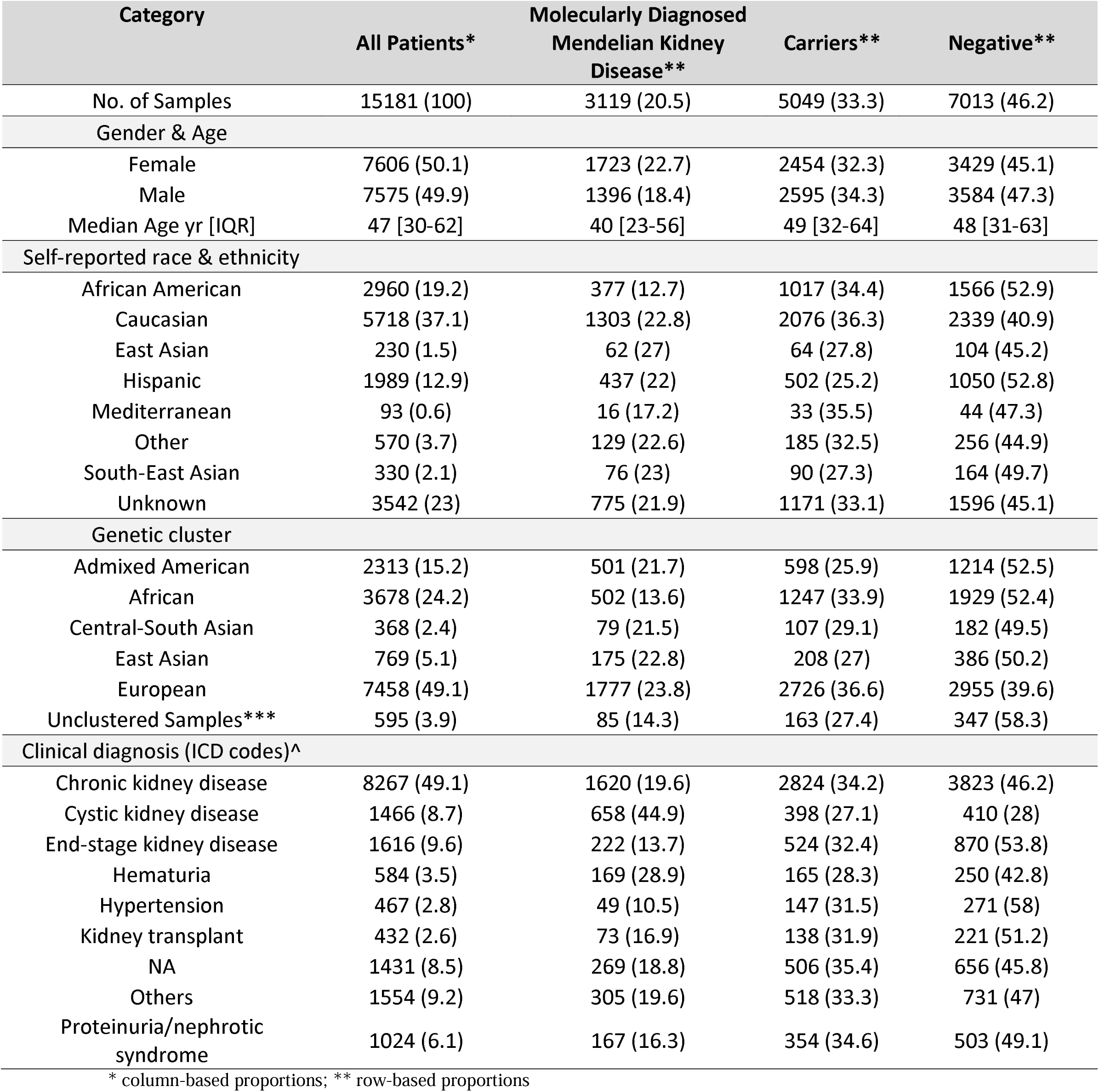
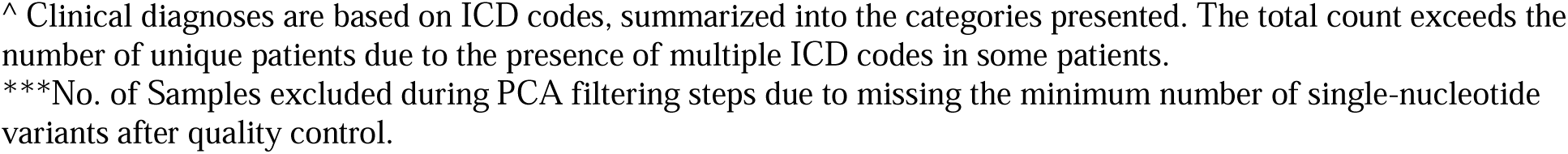
Clinical and demographic characteristics among patients, stratified by Mendelian kidney disease status.

| Category | All Patients* | Molecularly Diagnosed<br>Mendelian Kidney<br>Disease** | Carriers** | Negative** |
| --- | --- | --- | --- | --- |
| No. of Samples | 15181 (100) | 3119 (20.5) | 5049 (33.3) | 7013 (46.2) |
| Gender & Age |  |  |  |  |
| Female | 7606 (50.1) | 1723 (22.7) | 2454 (32.3) | 3429 (45.1) |
| Male | 7575 (49.9) | 1396 (18.4) | 2595 (34.3) | 3584 (47.3) |
| Median Age yr [IQR] | 47 [30-62] | 40 [23-56] | 49 [32-64] | 48 [31-63] |
| Self-reported race & ethnicity |  |  |  |  |
| African American | 2960 (19.2) | 377 (12.7) | 1017 (34.4) | 1566 (52.9) |
| Caucasian | 5718 (37.1) | 1303 (22.8) | 2076 (36.3) | 2339 (40.9) |
| East Asian | 230 (1.5) | 62 (27) | 64 (27.8) | 104 (45.2) |
| Hispanic | 1989 (12.9) | 437 (22) | 502 (25.2) | 1050 (52.8) |
| Mediterranean | 93 (0.6) | 16 (17.2) | 33 (35.5) | 44 (47.3) |
| Other | 570 (3.7) | 129 (22.6) | 185 (32.5) | 256 (44.9) |
| South-East Asian | 330 (2.1) | 76 (23) | 90 (27.3) | 164 (49.7) |
| Unknown | 3542 (23) | 775 (21.9) | 1171 (33.1) | 1596 (45.1) |
| Genetic cluster |  |  |  |  |
| Admixed American | 2313 (15.2) | 501 (21.7) | 598 (25.9) | 1214 (52.5) |
| African | 3678 (24.2) | 502 (13.6) | 1247 (33.9) | 1929 (52.4) |
| Central-South Asian | 368 (2.4) | 79 (21.5) | 107 (29.1) | 182 (49.5) |
| East Asian | 769 (5.1) | 175 (22.8) | 208 (27) | 386 (50.2) |
| European | 7458 (49.1) | 1777 (23.8) | 2726 (36.6) | 2955 (39.6) |
| Unclustered Samples*** | 595 (3.9) | 85 (14.3) | 163 (27.4) | 347 (58.3) |
| Clinical diagnosis (ICD codes)^ |  |  |  |  |
| Chronic kidney disease | 8267 (49.1) | 1620 (19.6) | 2824 (34.2) | 3823 (46.2) |
| Cystic kidney disease | 1466 (8.7) | 658 (44.9) | 398 (27.1) | 410 (28) |
| End-stage kidney disease | 1616 (9.6) | 222 (13.7) | 524 (32.4) | 870 (53.8) |
| Hematuria | 584 (3.5) | 169 (28.9) | 165 (28.3) | 250 (42.8) |
| Hypertension | 467 (2.8) | 49 (10.5) | 147 (31.5) | 271 (58) |
| Kidney transplant | 432 (2.6) | 73 (16.9) | 138 (31.9) | 221 (51.2) |
| NA | 1431 (8.5) | 269 (18.8) | 506 (35.4) | 656 (45.8) |
| Others | 1554 (9.2) | 305 (19.6) | 518 (33.3) | 731 (47) |
| Proteinuria/nephrotic syndrome | 1024 (6.1) | 167 (16.3) | 354 (34.6) | 503 (49.1) |

\* column-based proportions; \*\* row-based proportions
^ Clinical diagnoses are based on ICD codes, summarized into the categories presented. The total count exceeds the number of unique patients due to the presence of multiple ICD codes in some patients.
\*\*\*No. of Samples excluded during PCA filtering steps due to missing the minimum number of single-nucleotide variants after quality control.

### Prevalence of Mendelian kidney disease in the overall cohort

Of 15,181 patients, 3119 had a molecular diagnosis of Mendelian kidney disease (20.5%; **Table 1**), defined by the presence of pathogenic or likely pathogenic variants (P/LP) consistent with the inheritance pattern of the specific disease. 5049 (33.3%) patients were classified as carriers of Mendelian kidney disease, and 7013 (46.2%) were negative for P/LP variants in any of the 343 genes evaluated (**Supplemental Table 1**). Patients with a molecular diagnosis of Mendelian kidney disease were younger on average, at the time of testing, compared to carriers or patients with negative test results (40 v. 48 years, p<0.0005; **Table 1**).

We grouped Mendelian forms of kidney disease into five non-exclusive categories: (1) cystic and tubulointerstitial diseases, (2) glomerular diseases, (3) tubulopathies and other tubular diseases, (4) congenital anomalies of the kidney and urinary tract (CAKUT) and other structural disorders, and (5) complement-related kidney diseases (see **Supplemental Table 1**). Of patients with a molecular diagnosis of Mendelian kidney disease, 42.1% had cystic and tubulointerstitial disorders (dominated by variants in *PKD1/2*), 34.4% had glomerular disorders (dominated by variants in *COL4A3/4/5*), 13.9% tubulopathies and other tubular disorders, 8.0% had CAKUT and other structural disorders and 1.5% had complement-related diseases. In total (including molecular diagnosis positive and carriers), 4820 rare P or LP variants were identified across 286 of the 343 genes evaluated. Nearly half of these variants (46.8%, n=2,258) were not present in ClinVar or gnomAD databases at the time of the variants being assessed. The reported variants were categorized as frameshift (28%, n=1352), missense (24.3%, n=1170), nonsense (21.4%, n=1032), canonical splice site (14.5%, n=700), non-frameshift insertions/deletions (5%, n=240), copy number (CNV, 3.4%, n=166), or other (∼3.2%, n=160). The majority of patients with a molecular diagnosis of Mendelian kidney diseases (71.5%; n=2334) had variants in one (or more) of 10 genes: *PKD1/2, COL4A3/4/5, HNF1B, SLC12A3, UMOD, GLA,* and *SLC7A9* (Supplemental Table 5).

### Prevalence of APOL1 risk alleles

Of 15,181 individuals, 1035 (6.8%) had high-risk *APOL1* genotypes (**Table 2**). Homozygous haplotypes G1/G1 and G2/G2 accounted for 43.7% (n = 452) and 12.9% (n = 134) of patients, respectively; 449 patients with high-risk *APOL1* genotypes had one G1 allele and one G2 allele (43.4%; G1/G2). 1687 (11.1%) patients were single-risk APOL1 allele carriers, of which 1064 with G1 (63.1%), and 623 with G2 (36.9%); 12459 patients were G0/G0 (82.1%).

**Table 2.** Demographic data of patients included, stratified by presence of *APOL1* risk alleles.

| Category | All Patients | High-Risk *APOL1* Genotype |  |  |  | Single *APOL1* Risk Allele Carriers |  |  |  | G0/G0 Genotypes |  |  |  |
| --- | --- | --- | --- | --- | --- | --- | --- | --- | --- | --- | --- | --- | --- |
|  |  | All | Positive* | Carrier | Negative | All | Positive* | Carrier | Negative | All | Positive* | Carrier | Negative |
| No. of Samples | 15181 | 1035 | 95 | 367 | 573 | 1687 | 243 | 563 | 881 | 12459 | 2781 | 4119 | 5559 |
| Gender & Age |  |  |  |  |  |  |  |  |  |  |  |  |  |
| Female | 7606 | 472 | 54 | 152 | 266 | 798 | 133 | 248 | 417 | 6336 | 1536 | 2054 | 2746 |
| Male | 7575 | 563 | 41 | 215 | 307 | 889 | 110 | 315 | 464 | 6123 | 1245 | 2065 | 2813 |
| Median Age yr [IQR] | 47 [30-62] | 45 [32-57] | 37 [22-52] | 46 [35-57] | 45 [32-57] | 46 [31-60] | 40 [22-53] | 47 [31-63] | 48 [33-61] | 47 [29-63] | 40 [24-57] | 50 [32-65] | 48 [31-64] |
| Self-reported race & ethnicity |  |  |  |  |  |  |  |  |  |  |  |  |  |
| African American | 2960 | 815 | 73 | 293 | 449 | 1158 | 154 | 404 | 600 | 987 | 150 | 320 | 517 |
| Caucasian | 5718 | 9 | 1 | 3 | 5 | 47 | 13 | 12 | 22 | 5662 | 1289 | 2061 | 2312 |
| East Asian | 230 | 0 | 0 | 0 | 0 | 0 | 0 | 0 | 0 | 230 | 62 | 64 | 104 |
| Hispanic | 1989 | 41 | 2 | 17 | 22 | 136 | 26 | 39 | 71 | 1812 | 409 | 446 | 957 |
| Mediterranean | 93 | 0 | 0 | 0 | 0 | 1 | 0 | 0 | 1 | 92 | 16 | 33 | 43 |
| Other | 570 | 23 | 2 | 12 | 9 | 25 | 4 | 11 | 10 | 522 | 123 | 162 | 237 |
| South-East Asian | 330 | 0 | 0 | 0 | 0 | 2 | 0 | 0 | 2 | 328 | 76 | 90 | 162 |
| Unknown | 3542 | 161 | 18 | 50 | 93 | 341 | 50 | 107 | 184 | 3040 | 707 | 1014 | 1319 |
| Genetic cluster |  |  |  |  |  |  |  |  |  |  |  |  |  |
| Admixed American | 2313 | 27 | 0 | 8 | 19 | 141 | 25 | 38 | 78 | 2145 | 476 | 552 | 1117 |
| African | 3678 | 944 | 91 | 345 | 508 | 1453 | 198 | 496 | 759 | 1281 | 213 | 406 | 662 |
| Central-South Asian | 368 | 0 | 0 | 0 | 0 | 2 | 1 | 0 | 1 | 366 | 78 | 107 | 181 |
| East Asian | 769 | 0 | 0 | 0 | 0 | 1 | 0 | 1 | 0 | 768 | 175 | 207 | 386 |
| European | 7458 | 4 | 1 | 1 | 2 | 37 | 10 | 10 | 17 | 7417 | 1766 | 2715 | 2936 |
| Unclustered Samples*** | 595 | 60 | 3 | 13 | 44 | 53 | 9 | 18 | 26 | 482 | 73 | 132 | 277 |

\*Positive refers to patients with a molecular diagnosis of Mendelian kidney disease
\*\*\*No. of Samples excluded during PCA filtering steps due to missing the minimum number of single-nucleotide variants after quality control.

The large majority of patients with high-risk *APOL1* genotypes had designated African American race (815/1035; 78.9%). Similarly, single-risk allele carriers had a high proportion of designated African American race (1158/1687; 68.6%), in contrast to individuals with G0/G0 genotypes, who had a low proportion of designated African American race (987/12459; 7.9%). We chose to deliberately study patients with single-risk *APOL1* alleles, despite being low-risk, because, by definition, these have recent African ancestry, and as such make a relevant comparator group to patients with high-risk *APOL1* genotypes. Though ∼23% of patients in our cohort had unknown designated race/ethnicity information, we were able to recategorize these patients according to their genomic ancestry clusters. Among patients with high-risk *APOL1* genotypes and unknown designated race/ethnicity, the vast majority (94%) were classified as having recent African ancestry (**Supplemental Figure 2** and **Supplemental Table 6**). Finally, 25.6% of patients in the genetic African ancestry cluster had high-risk *APOL1* genotypes (n= 944/3678; **Table 2**).

### Prevalence of Mendelian kidney disease stratified by APOL1 genotype and recent African ancestry

Of 1035 patients with high-risk *APOL1* genotypes, 95 (9.2%) had a concomitant molecular diagnosis of Mendelian kidney disease (**Table 2**), a smaller fraction compared to single-risk *APOL1* allele carriers (14.4%, p<0.0001), and patients with G0/G0 (22.3%, p<0.00001). The most common forms of Mendelian kidney disease among patients with high-risk *APOL1* alleles were: Alport syndrome/thin basement membrane disease (3.7%), autosomal dominant polycystic kidney disease (ADPKD; 2.0%), and *HNF1B*-associated disease (0.5%). The full list of molecularly diagnosed Mendelian kidney disease are presented in **Supplemental Table 5**.

The overall prevalence of molecularly diagnosed Mendelian kidney disease was significantly lower among patients with recent genetic African ancestry compared to those with recent European ancestry (13.6% v. 23.8%, p<0.00001). Even among patients with G0/G0 genotypes, those with recent genetic African ancestry had a lower prevalence of molecular diagnosed Mendelian kidney disease compared to patients with European ancestry (16.6% v. 23.8%, p<0.00001). To evaluate the possibility that recent African ancestry might confound the association between *APOL1* risk alleles and molecularly diagnosed Mendelian kidney disease, we matched patients with high-risk *APOL1* genotypes to single-risk allele carriers and patients with G0/G0, in a 1:1 ratio, based on age, sex, the genomic proportions of continental populations, and the first four principal components (**Supplemental Table 7**). We found a significant lower prevalence of molecularly diagnosed Mendelian kidney disease among patients with high-risk *APOL1* risk alleles (9.4%; n=92) compared to matched single-risk allele carriers (13.4%; n=128, p=0.006) and matched G0/G0 patients (16.2%; n=156, p=0.000009). We did not observe statistically significant difference in the prevalence of molecularly diagnosed Mendelian kidney disease between matched single-risk allele carriers and matched G0/G0 patients (p=0.09). These finding suggest that the lower prevalence of molecularly diagnosed Mendelian kidney disease among patients with high-risk *APOL1* genotypes is not driven by differences in the proportions of patients with recent African ancestry across the *APOL1* genotypes.

## DISCUSSION

Because the lifetime risk of developing kidney failure among persons with high-risk *APOL1* genotypes is ∼15%, there is a need to understand how other genetic and non-genetic modifiers contribute to the risk of kidney disease and kidney disease progression (19). Here we studied Mendelian kidney disease in individuals with high-risk *APOL1* genotypes using genomic data derived from a multi-ethnic cohort of more than 15,000 patients who underwent commercial clinical genetic testing for Mendelian kidney disease in the United States.

We found that 9.2% of patients with high-risk *APOL1* genotypes in this cohort had concurrent molecularly diagnosed Mendelian kidney disorders. To our knowledge, only one single-center study (n=239 patients with high-risk *APOL1* genotypes) has explored the prevalence of Mendelian kidney disease among patients with CKD and *APOL1* high-risk genotypes and CKD (19), demonstrating that 2.5% of these patients had concurrent Mendelian kidney diseases. In contrast, our cohort of patients undergoing commercial clinical genetic testing had a much higher prevalence. While the difference in prevalence estimates is likely explained by more refined patient selection in our cohort of patients undergoing clinical genetic testing, it also highlights the potential contribution of Mendelian forms of kidney disease among patients with high-risk *APOL1* genotypes. The majority of patients with high-risk *APOL1* genotypes and concurrent molecularly diagnosed Mendelian kidney disorders in our cohort had glomerular disorders (44.9%), dominated by variants in Alport syndrome genes: *COL4A3*, *COL4A4* and *COL4A5*. Because patients with *APOL1*-mediated kidney disease and Alport syndrome can present with overlapping clinical (e.g., hypertension, proteinuria) and histologic findings on kidney biopsy (e.g. focal segmental glomerulosclerosis), this finding may support consideration of diagnostic genetic testing in a subset of patients with high-risk *APOL1* genotypes.

Our analysis also demonstrated that patients with recent African ancestry had lower prevalence of molecularly diagnosed Mendelian kidney disease than patients with more remote African Ancestry or patients without African ancestry. This finding aligns with existing research indicating a generally lower diagnostic yield for genetic diseases in African-American patients with kidney disease (20–22). Such discrepancies might be attributed to the underrepresentation of African Americans in genomic sequencing studies (22–24). This work highlights the potential need for more equitable genetic disease identification across racial and ethnic groups, including African Americans, a critical area for future research.

Several limitations in this study should be acknowledged. First, our cohort is composed of a selected population of patients undergoing clinical genetic testing for Mendelian kidney disease; we had limited clinical information, and we can only infer (but not document) that these patients had kidney disease and that a genetic cause was contemplated. As such, we should be cautious in extrapolating these findings to unrestricted populations of patients with CKD. The lack of clinical information prevents genotype-phenotype correlation, and as such, we have used the term “molecularly diagnosed Mendelian kidney disease,” as we have no information on any patient’s phenotype. Despite this limitation, we note that variants classified as P/LP meet stringent criteria, and in a patient, population selected for clinical genetic testing, the likelihood that such variants contribute causally to a patient’s kidney disease would be high. Second, our analysis focused on a set of 343 genes known to be associated with kidney disease, which, while extensive, may not encompass all possible genetic variants contributing to kidney disease.

Overall, our data suggest that Mendelian kidney disease could contribute to the development and progression of CKD in a fraction of patients with high-risk *APOL1* genotypes. Future research to confirm these findings in diverse, unselected cohorts, and further elucidate the interaction between Mendelian kidney disease and *APOL1* risk alleles is needed.

## Data Availability

Partial restrictions to the data and/or materials apply. All available data is included in the manuscript and/or supporting information.

## Acknowledgements

V.C. is supported by KL2 TR003143.

## Guarantor

VC takes full responsibility for the work, including the study design, access to data, and the decision to submit and publish the manuscript.

## Authors’ Contributions

RSFJ: Research idea and study design, data analysis and interpretation, statistical analysis

SP: Research idea and study design, data analysis and interpretation

LV: Research idea and study design, data analysis and interpretation

NS: Research idea and study design, data analysis and interpretation

VB: Data interpretation

GMC: Data interpretation

DKK: Research idea and study design, data analysis and interpretation

VC: Research idea and study design, data analysis and interpretation, statistical analysis

Each author contributed important intellectual content during manuscript drafting or revision and agrees to be personally accountable for the individual’s own contributions and to ensure that questions pertaining to the accuracy or integrity of any portion of the work, even one in which the author was not directly involved, are appropriately investigated and resolved, including with documentation in the literature if appropriate.

