## Supplementary information for "Prevalence of Mendelian kidney disease among patients with high-risk *APOL1* genotypes undergoing commercial genetic testing in the United States"

**Supplemental Figure 1:** Comparison of self-reported/designated race/ethnicity with genomic ancestry derived from HGDP and 1KG.

**Supplemental Figure 2:** Population Stratification in 1KGP and HGDP Samples highlighting *APOL1* Allele Carriers.

**Supplemental Table 1:** List of 343 Genes Included in our Study.

**Supplemental Table 2:** Comparative Analysis of Self-Reported/Designated Race/Ethnicity and Genetic Clusters Derived from Principal Component Analysis.

**Supplemental Table 3:** Distribution of Global Ancestry Proportions Across Five Continental Populations in Genetic Clusters from Principal Component Analysis.

**Supplemental Table 4:** Global Ancestry Proportions in Five Continental Populations Based on Self-Reported Race and Ethnicity.

**Supplemental Table 5:** Frequency of molecular diagnosed kidney disease across the 343 genes studied, stratified by *APOL1* Genotype.

**Supplemental Table 6:** Detailed Characteristics of Samples with *APOL1* Risk Alleles

**Supplemental Table 7:** Detailed Characteristics of Samples Selected by Matching Analysis

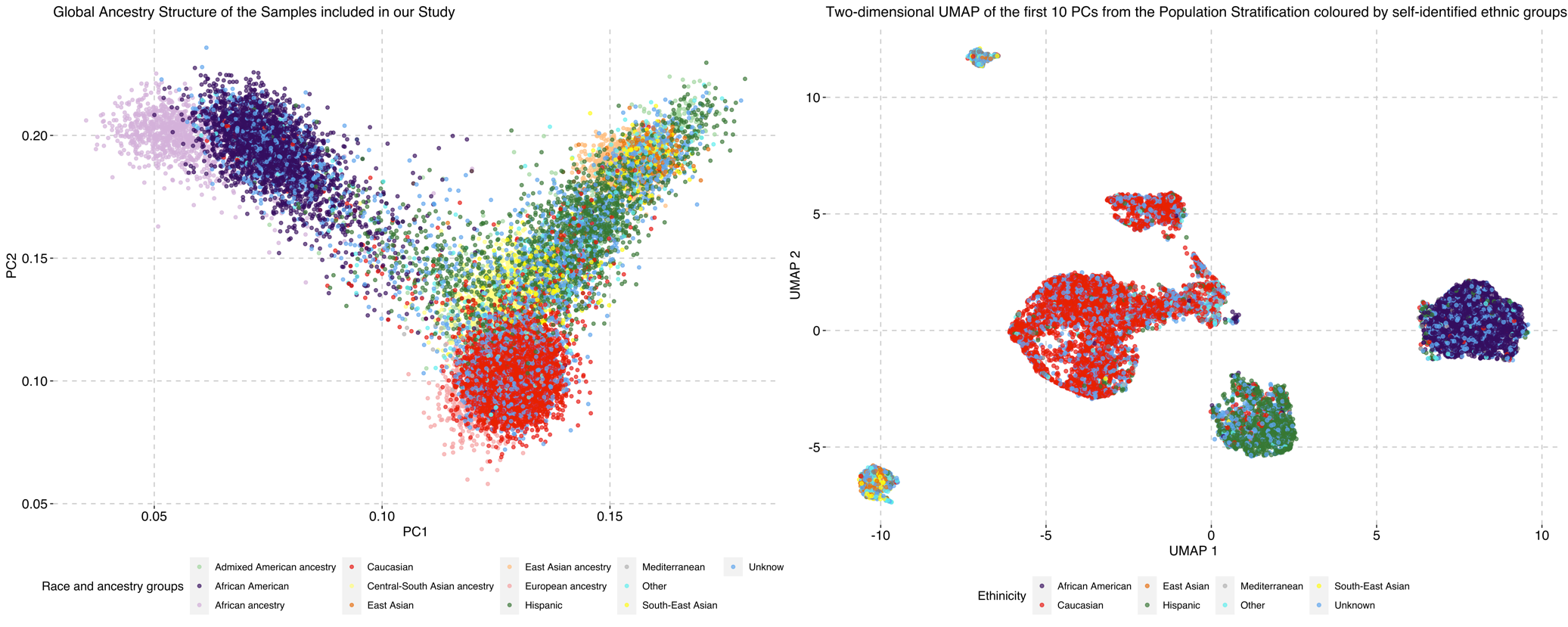

**Supplemental Figure 1: Comparison of self-reported or designated race/ethnicity with genomic ancestry derived from HGDP and 1KG.** *Left Panel:* Each point represents an individual. Points in transparent colors demonstrate individuals from HGDP and 1KG, and points in darker (opaque) colors demonstrate the self-reported race/ethnicity from individuals from our cohort. PC1 and PC2 represent the first two principal components of ancestry informative markers. The genetic diversity across the samples includes five major continental populations: European, African, Admixed American, Central-South Asian, and East Asian, alongside the self-reported race/ethnicity of participants in our study. There is excellent qualitative overlap between the self-reported race/ethnicity and the expected genetic ancestry. *Right Panel:* UMAP visualization based on the first 10 principal components. The sample coloring corresponds to the participants' self-reported race/ethnicity, providing insights into the clustering patterns relative to self-identified groups.

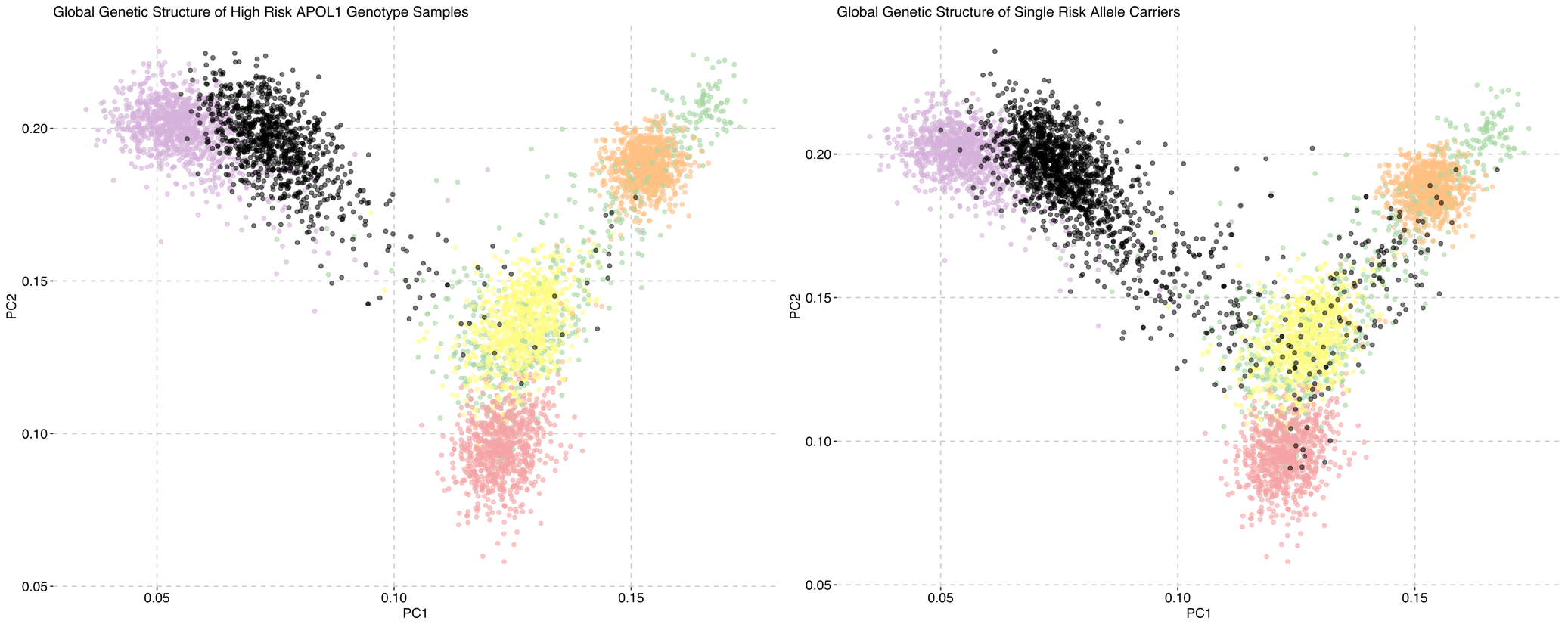

**Supplemental Figure 2: Population Stratification in 1KGP and HGDP Samples highlighting *APOL1* Allele Carriers.** *Left Panel:* This panel shows the global ancestry structure of individuals with high-risk *APOL1* genotypes, represented by black points, illustrating their distribution across the diverse population groups in 1KGP and HGDP Samples. *Right Panel:* Single risk allele carriers for *APOL1* are represented by black points. It is clear in both panels that individuals with high-risk *APOL1* genotypes and single risk allele carriers have recent African ancestry (transparent purple points).

**Supplemental Table 1: List of 343 genes and Mendelian conditions studied.**

| **Gene** | **Disease Name** | **Inheritance** | **Disease Category** | **P/LP variants** |
| --- | --- | --- | --- | --- |
| *ABCC6* | Pseudoxanthoma elasticum and/or Generalized arterial calcification of infancy | AR | T | Yes |
| *ABCC8* | ABCC8-related disorders | AD;AR | T; G | Yes |
| *ACE* | Renal Tubular Dysgenesis | AR | CS | Yes |
| *ACTB* | ACTB-Related Disorders | AD | CS | No |
| *ACTN4* | Focal Segmental Glomerulosclerosis | AD |  | Yes |
| *ADA2* | Adenosine Deaminase 2 Deficiency (DADA2); Sneddon syndrome | AR | G | Yes |
| *ADAMTS13* | Familial Thrombotic Thrombocytopenic Purpura | AR | CR | Yes |
| *AGPAT2* | Lipodystrophy congenital generalized, type 1 | AR | G | Yes |
| *AGT* | Renal Tubular Dysgenesis | AR | CS | Yes |
| *AGTR1* | Renal Tubular Dysgenesis | AR | CS | Yes |
| *AGXT* | Primary hyperoxaluria type 1 | AR | T | Yes |
| *AHI1* | Joubert Syndrome 3 | AR | CTI | Yes |
| *ALG1* | Congenital Disorder of Glycosylation, Type 1K | AR | G | Yes |
| *ALG8* | Congenital disorder of glycosylation type Ih | AR | CTI | Yes |
| *ALG9* | Polycystic Kidney and Liver Disease (AD); Congenital Disorder of Glycosylation, type 2 (AR); Gillessen Kaesbach Nishimura syndrome (GIKANIS) (AR) | AD;AR | CTI | Yes |
| *ALMS1* | Alstrom syndrome | AR | CTI | Yes |
| *ALPL* | Hypophosphatasia | AD;AR | T | Yes |
| *AMN* | Megaloblastic Anemia 1, Norwegian Type | AR | T | Yes |
| *ANKS6* | Nephronophthisis 16 | AR | CTI | Yes |
| *ANOS1* | Isolated Gonadotropin-Releasing Hormone (GnRH) Deficiency | XL | CS | Yes |
| *AP2S1* | Familial Hypocalciuric Hypercalcemia, Type 3 | AD | T | No |
| *APOA1* | Familial Visceral Amyloidosis (AD); Primary Hypoalphalipoproteinemia (AR) | AD;AR | G | Yes |
| *APOC2* | Hyperlipoproteinemia, type Ib | AR | G | Yes |
| *APOPT1* | Mitochondrial Complex 4 deficiency | AR | T | Yes |
| *APRT* | Adenine Phosphoribosyltransferase Deficiency | AR | T | Yes |
| *AQP2* | Diabetes insipidus, nephrogenic | AD;AR | T | Yes |
| *ARL6* | Bardet-Biedl Syndrome 3 (AR); Retinitis Pigmentosa 55 (AR) | AR | CTI | Yes |
| *ATP6V0A4* | ATP6V0A4-Distal Renal Tubular Acidosis (AR) | AR | T | Yes |
| *ATP6V1B1* | ATP6V1B1-Distal Renal Tubular Acidosis (ATP6V1B1-dRTA) | AR | T | Yes |
| *ATP7B* | Wilson disease | AR | T | Yes |
| *AVP* | Diabetes insipidus, Neurohypophyseal | AD | T | Yes |
| *AVPR2* | Diabetes Insipidus, Nephrogenic | XL | T | Yes |
| *B2M* | Familial Visceral Amyloidosis (AD); Immunodeficiency 43 (AR) | AD;AR | G | Yes |
| *BBS1* | Bardet-Biedl syndrome 1 | AR | CS; CTI | Yes |
| *BBS10* | Bardet-Biedl syndrome 10 | AR | CS; CTI | Yes |
| *BBS12* | Bardet-Biedl syndrome 12 | AR | CS; CTI | Yes |
| *BBS2* | Bardet-Biedl syndrome 2 (AR); Retinitis Pigmentosa 74 (AR) | AR | CS; CTI | Yes |
| *BBS4* | Bardet-Biedl Syndrome 4 | AR | CS; CTI | Yes |
| *BBS5* | Bardet-Biedl Syndrome 5 | AR | CS; CTI | Yes |
| *BBS7* | Bardet-Biedl syndrome 7 | AR | CS; CTI | Yes |
| *BBS9* | Bardet-Biedl syndrome 9 | AR | CTI | Yes |
| *BCS1L* | BCS1L-Related Disorders (AR) | AR | T | Yes |
| *BICC1* | Renal Dysplasia, Cystic | UK | CS; CTI | No |
| *BMP4* | Congenital Anomalies of the Kidney and Urinary Tract; Microphthalmia, syndromic 6 | AD | CS | No |
| *BMPR2* | Pulmonary Hypertension, Familial Primary with or without Hereditary Hemorrhagic Telangiectasia & Pulmonary venoocclusive disease 1 | AD | T | Yes |
| *BRAF* | Noonan Syndrome (AD); Noonan Syndrome with Multiple Lentigines (AD); Cardiofaciocutaneous Syndrome (AD) | AD | CS | No |
| *BSCL2* | BSCL2-related conditions | AD;AR | G | Yes |
| *BSND* | Bartter Syndrome, Type 4a (AR) | AR | T | Yes |
| *C3* | Atypical Hemolytic Uremic Syndrome (AD); C3 Glomerulopathy (AD); C3 deficiency (AR) | AD;AR | CR | Yes |
| *CA2* | Osteopetrosis with Renal Tubular Acidosis | AR | T | Yes |
| *CACNA1H* | Familial Hyperaldosteronism, Type IV (AD) | AD | T | No |
| *CACNA1S* | Hypokalemic Periodic Paralysis Type 1 (AD); Malignant Hyperthermia Susceptibility (AD); Dihydropyridine receptor congenital myopathy (AR) | AD;AR |  | Yes |
| *CASR* | CASR-related conditions | AD;AR | T | Yes |
| *CAV1* | Lipodystrophy, Familial Partial, type 7 (AD); Pulmonary Hypertension, Primary, 3 (AD); Lipodystrophy, Congenital Generalized, Type 3 (AR) | AD;AR | T | Yes |
| *CD151* | Nephropathy with Pretibial Epidermolysis Bullosa and Deafness | AR | G | Yes |
| *CD2AP* | Focal Segmental Glomerulosclerosis 3 (AR); Susceptibility to Focal Segmental Glomerulosclerosis (AD) | AD;AR | G | Yes |
| *CDC73* | CDC73-related Disorders (AD) | AD | T | Yes |
| *CDKN1C* | Beckwith-Wiedemann Syndrome (AD); IMAGe Syndrome (AD) | AD | T | No |
| *CEL* | Maturity-Onset Diabetes of the Young, Type 8 | AD | G | Yes |
| *CEP164* | Senior-Loken Syndrome; Nephronophthisis 15 | AR | CTI | Yes |
| *CEP290* | CEP290-Related Ciliopathies | AR | CTI | Yes |
| *CFH* | Atypical Hemolytic Uremic Syndrome (AD); C3 Glomerulopathy (AD); Complement Factor H Deficiency (AR) | AD;AR | CR | Yes |
| *CFHR5* | Atypical Hemolytic Uremic Syndrome (AD); C3 Glomerulopathy (AD); Complement Factor H Deficiency (AR) | AD;AR | CR | No |
| *CFI* | Atypical Hemolytic Uremic Syndrome (AD); C3 Glomerulopathy (AD); Complement Factor I Deficiency (AR) | AD;AR | CR | Yes |
| *CHD7* | CHARGE syndrome | AD | CS | Yes |
| *CHRM3* | Prune-Belly Syndrome (AR) | AR | CS | No |
| *CHRNA3* | Bladder Dysfunction, Autonomic, with Impaired Pupillary Reflex and Secondary Congenital Anomalies of the Kidney and Urinary Tract | AR | CS | Yes |
| *CISD2* | Wolfram Syndrome 2 | AR | G | No |
| *CLCN2* | Hyperaldosteronism (AD); CLCN2-related leukoencephalopathy (AR) | AD;AR | T | Yes |
| *CLCN5* | Dent disease 1 | XL | T | Yes |
| *CLCNKB* | Bartter Syndrome, Type 3/4B (AR); Gitelman syndrome (AR) | AR | T | Yes |
| *CLDN16* | Hypomagnesemia 3, Renal | AR | T | Yes |
| *CLDN19* | Hypomagnesemia 5, Renal, with Ocular Involvement | AR | T | Yes |
| *CNNM2* | Renal hypomagnesemia (AD/AR) | AD;AR | T | Yes |
| *COL4A1* | COL4A1-Related Disorders | AD | CTI | Yes |
| *COL4A3* | COL4A3-related Alport syndrome | AD;AR | G | Yes |
| *COL4A4* | COL4A4-related Alport syndrome | AD;AR | G | Yes |
| *COL4A5* | Alport syndrome, X-linked | XL | G | Yes |
| *COQ2* | Coenzyme Q10 Deficiency, Primary 1 | AR | G | Yes |
| *COQ6* | Coenzyme Q10 Deficiency, Primary 6 | AR | G | Yes |
| *COX10* | Mitochondrial Complex 4 deficiency | AR | T | Yes |
| *COX20* | Mitochondrial Complex 4 deficiency | AR | T | Yes |
| *COX6B1* | Mitochondrial Complex 4 deficiency | AR | T | No |
| *CPLANE1* | Orofaciodigital Syndrome 6 (AR); Joubert syndrome with Oral-Facial-Digital features (AR) | AR | CTI | Yes |
| *CPT2* | Carnitine Palmitoyltransferase II Deficiency (AR) | AR | G; T | Yes |
| *CREBBP* | Rubinstein-Taybi Syndrome, Type 1 (AD); Menke-Hennekam Syndrome (AD) | AD | T | No |
| *CTNS* | Cystinosis | AR | CTI | Yes |
| *CUBN* | Proteinuria, chronic benign; Megaloblastic Anemia 1, FinnishType | AR | G | Yes |
| *CUL3* | Pseudohypoaldosteronism, Type 2E | AD | T | Yes |
| *CYP11A1* | Congenital adrenal insufficiency (CYP11A1-deficiency) | AR | T | Yes |
| *CYP11B1* | Familial hyperaldosteronism type I (AD); Adrenal hyperplasia, congenital, due to 11-beta-hydroxylase deficiency (AR) | AD;AR | T | Yes |
| *CYP11B2* | Corticosterone methyloxidase deficiency | AR | T | Yes |
| *CYP17A1* | 17-Alpha-Hydroxylase/7,20-Lyase Deficiency | AR | T | Yes |
| *CYP24A1* | CYP24A1-related hypercalcemia (AR) | AR | T | Yes |
| *CYP27B1* | Vitamin D-Dependent Rickets, Type 1A | AR | T | Yes |
| *CYP2R1* | Rickets due to Defect in Vitamin D 25-hydroxylation | AR | T | Yes |
| *DCDC2* | Nephronophthisis 19 | AR | CTI | Yes |
| *DGKE* | Atypical Hemolytic Uremic Syndrome | AR | G | Yes |
| *DHCR7* | Smith-Lemli-Opitz syndrome | AR | CS | Yes |
| *DLC1* | Nephrotic Syndrome | UK | G | No |
| *DMP1* | Hypophosphatemic Rickets | AR | T | Yes |
| *DNASE1L3* | Systemic Lupus Erythematosus 16 | AR | T | Yes |
| *EBP* | Chondrodysplasia Punctata (XL); MEND syndrome (XL) | XL | T | No |
| *EIF2AK3* | Wolcott-Rallison Syndrome | AR | G | Yes |
| *ELP1* | Familial Dysautonomia, Hereditary Sensory and Autonomic Neuropathy Type 3 | AR | G | Yes |
| *ENPP1* | Hypophosphatemic Rickets (AR); Generalized arterial calcification of infancy (AR); Cole disease (AD) | AD;AR | T | Yes |
| *EYA1* | Branchiootorenal Spectrum Disorders (AD) | AD | CS | Yes |
| *FAM20A* | Amelogenesis Imperfecta, Type 1G | AR | T | Yes |
| *FAN1* | Interstitial Nephritis, Karyomegalic | AR | CTI | Yes |
| *FANCA* | Fanconi Anemia, Group A (AR) | AR | CS | Yes |
| *FANCB* | Fanconi Anemia, Group B | XL | CS | No |
| *FANCC* | Fanconi Anemia, Group C (AR) | AR | CS | Yes |
| *FANCD2* | Fanconi Anemia, Group D2 | AR | CS | Yes |
| *FANCE* | Fanconi Anemia, Group E | AR | CS | Yes |
| *FANCF* | Fanconi Anemia, Group F | AR | CS | Yes |
| *FANCG* | Fanconi Anemia, Group G | AR | CS | Yes |
| *FANCI* | Fanconi Anemia, Group I | AR | CS | Yes |
| *FANCL* | Fanconi Anemia, Group L | AR | CS | Yes |
| *FASTKD2* | Mitochondrial Complex 4 deficiency | AR | T | Yes |
| *FGA* | Hereditary Renal Amyloidosis (AD); Congenital Fibrinogen Deficiency (AR) | AD;AR | G | Yes |
| *FGF10* | Lacrimo-auriculo-dento-digital (LADD) Syndrome | AD | CS | No |
| *FGF23* | Hypophosphatemic Rickets (AD); Tumoral Calcinosis, Hyperphosphatemic (AR) | AD;AR | T | Yes |
| *FGFR1* | FGFR1-related conditions | AD | CS | Yes |
| *FGFR2* | FGFR2-related conditions | AD | CS | Yes |
| *FLCN* | Birt-Hogg-Dubé syndrome (AD); Primary Spontaneous Pneumothorax (AD) | AD | CTI | Yes |
| *FN1* | Glomerulopathy With Fibronectin Deposits 2 (AD); Corner Fracture Type Spondylometaphyseal Dysplasia (AD) | AD | G | Yes |
| *FOXC1* | Axenfeld-Rieger Syndrome, Type 3 | AD | CS | Yes |
| *FOXC2* | Lymphedema-Distichiasis Syndrome with Renal Disease and Diabetes Mellitus | AD | G | Yes |
| *FOXI1* | Hereditary Distal Renal Tubular Acidosis (AR) | AR | T | No |
| *FOXP3* | IPEX syndrome (including type 1 diabetes) | XL | G | Yes |
| *FRAS1* | Fraser Syndrome | AR | CS | Yes |
| *FREM1* | FREM1-related conditions | AD;AR | CS | Yes |
| *FREM2* | Fraser Syndrome | AR | CS | Yes |
| *FXYD2* | Hypomagnesemia 2, Renal | AD | T | No |
| *G6PC* | Glycogen storage disease, type 1a | AR | T | Yes |
| *GALNT3* | Hyperphosphatemic Familial Tumoral Calcinosis | AR | T | Yes |
| *GANAB* | Polycystic Kidney and/or Polycystic Liver Disease | AD | CTI | Yes |
| *GATA3* | Hypoparathyroidism, Sensorineural Deafness, and Renal Dysplasia | AD | CS | Yes |
| *GATM* | Cerebral creatine deficiency syndrome 3 (AR); Fanconi renotubular syndrome 1 (AD) | AD;AR | T | Yes |
| *GCK* | Maturity-onset diabetes of the young, type 2 (AD); Late onset, non insulin dependent diabetes mellitus (AD); Permanent neonatal diabetes mellitus 1 (AR); Familial hyperinsulinemic hypoglycemia 3 (AD) | AD;AR | G | Yes |
| *GCM2* | Familial Isolated Hypoparathyroidism (AD/AR); Familial Isolated Hyperparathyroidism (AD) | AD;AR | T | Yes |
| *GLA* | Fabry disease | XL | G | Yes |
| *GLI3* | GLI3-Related Disorders | AD | CS | Yes |
| *GLIS2* | Nephronophthisis 7 | AR | CTI | No |
| *GLIS3* | Diabetes Mellitus, Neonatal, With Congenital Hypothyroidism | AR | G; CTI | Yes |
| *GNA11* | Autosomal Dominant Hypocalcemia 2 (AD); Hypocalciuric hypercalcemia, type II (AD) | AD | T | No |
| *GNAS* | GNAS-Related Disorders | AD | T | Yes |
| *GPC3* | Simpson-Golabi-Behmel Syndrome, Type 1 | XL | CS | Yes |
| *GRHPR* | Hyperoxaluria, Primary, Type 2 | AR | T | Yes |
| *GRIP1* | Fraser Syndrome | AR | CS | Yes |
| *GSN* | Amyloidosis, Finnish Type | AD | G | Yes |
| *HBB* | Beta-Hemoglobinopathies (AD/AR); Sickle cell disease (AR) | AD;AR | G; T | Yes |
| *HGD* | Alkaptonuria | AR | T | Yes |
| *HNF1A* | HNF1A-MODY (Maturity-Onset Diabetes of the Young) | AD | CTI | Yes |
| *HNF1B* | HNF1B-Related Disorders | AD | CTI; CS | Yes |
| *HNF4A* | Fanconi Renotubular Syndrome 4, with or without Maturity-Onset Diabetes of the Young (MODY), Type 1 (AD); MODY, Type 1 (AD); Congenital Hyperinsulinism (AD); Diabetes Mellitus, Type 2 (AD) | AD | G | Yes |
| *HOGA1* | Primary Hyperoxaluria, Type 3 | AR | T | Yes |
| *HOXA13* | Hand-Foot-Uterus Syndrome | AD | CS | Yes |
| *HPRT1* | HPRT1-Related Disorders (XL) | XL | T | No |
| *HPS1* | Hermansky-Pudlak syndrome 1 | AR | G | Yes |
| *HPSE2* | Urofacial Syndrome (AR) | AR | CS | Yes |
| *HSD11B2* | Apparent Mineralocorticoid Excess | AR | T | Yes |
| *HSD3B2* | Congenital adrenal hyperplasia due to 3-beta-hydroxysteroid dehydrogenase 2 deficiency | AR | T | Yes |
| *IFT122* | Cranioectodermal Dysplasia, Type 1 | AR | CTI | Yes |
| *IFT140* | Retinitis pigmentosa 80; Short-Rib Thoracic Dysplasia 9 with or without Polydactyly | AR | CTI | Yes |
| *IFT172* | Short-Rib Thoracic Dysplasia 10 With Or Without Polydactyly (AR); Bardet-Biedl syndrome 20 (AR); Retinitis Pigmentosa 71 (AR) | AR | CTI | Yes |
| *IFT43* | Cranioectodermal Dysplasia, Type 3 | AR | CTI | Yes |
| *INF2* | Focal Segmental Glomerulosclerosis 5; Charcot-Marie-Tooth Disease E | AD | G | Yes |
| *INS* | Diabetes mellitus, insulin-dependent, 2 (AD); Diabetes mellitus, permanent neonatal 4 (AD/AR); Maturity-onset diabetes of the young, type 10 (AD); Hyperproinsulinemia (AD) | AD;AR | G | Yes |
| *INVS* | Nephronophthisis 2; Senior-Loken syndrome | AR | CTI | Yes |
| *IQCB1* | Senior-Loken Syndrome 5 | AR | CTI | Yes |
| *ITGA3* | Interstitial Lung Disease with Nephrotic Syndrome and Epidermolysis Bullosa | AR | G | Yes |
| *ITGA6* | Junctional Epidermolysis Bullosa- Pyloric Atresia Syndrome | AR | CS | Yes |
| *ITGB4* | Junctional Epidermolysis Bullosa-Pyloric Atresia Syndrome | AD;AR | G | Yes |
| *JAG1* | Alagille Syndrome, Type 1 | AD | CS | Yes |
| *KANSL1* | Koolen-De Vries Syndrome | AD | CS | Yes |
| *KAT6B* | Genitopatellar syndrome; SBBYSS syndrome | AD | CS | No |
| *KCNA1* | Episodic Ataxia Type 1 | AD | T | No |
| *KCNJ1* | Bartter syndrome | AR | T | Yes |
| *KCNJ10* | Seizures, Sensorineural Deafness, Ataxia, Mental Retardation, and Electrolyte Imbalance (SeSAME Syndrome) | AR | T | Yes |
| *KCNJ11* | KCNJ11-Related Diabetes (AD); Familial Hyperinsulinism (AD/AR)0 | AD;AR | G | Yes |
| *KCNJ5* | Familial Hyperaldosteronism Type 3 | AD | T | No |
| *KCNK3* | Pulmonary Hypertension, Primary 4 | AD | T | No |
| *KCTD1* | Scalp-Ear-Nipple Syndrome | AD | CS | No |
| *KLHL3* | Pseudohypoaldosteronism, Type 2D | AD;AR | T | Yes |
| *KRAS* | Cardiofaciocutaneous Syndrome (AD); Noonan Syndrome (AD) | AD | CS | No |
| *LAMB2* | Nephrotic Syndrome, Type 5, with or without Ocular Abnormalities; Pierson Syndrome | AR | G | Yes |
| *LCAT* | Complete LCAT Deficiency (AR); Partial LCAT Deficiency (AR) | AR | G | Yes |
| *LDHA* | Glycogen Storage Disease, Type 11 | AR | T | Yes |
| *LMNA* | LMNA-Related Disorders | AD;AR | G | Yes |
| *LMX1B* | Nail-Patella Syndrome (AD); Focal Segmental Glomerulosclerosis 10 (AD) | AD | G | Yes |
| *LPIN1* | Myoglobinuria, Acute Recurrent | AR | G | Yes |
| *LRP2* | Donnai-Barrow Syndrome | AR | T | Yes |
| *LRP4* | Cenani-Lenz Syndactyly Syndrome | AR | CS | Yes |
| *LRP5* | LRP5-Related Disorders | AD;AR | CTI | Yes |
| *LYZ* | Amyloidosis, Familial Visceral, Renal | AD | G | No |
| *LZTFL1* | Bardet-Biedl Syndrome, Type 17 | AR | CTI | Yes |
| *MAFB* | Multicentric Carpotarsal Osteolysis with or without Nephropathy | AD | G | Yes |
| *MAGI2* | Nephrotic Syndrome 15 | AR | G | Yes |
| *MEFV* | Familial Mediterranean fever | AR | G | Yes |
| *MKKS* | Bardet-Biedl Syndrome, Type 6 (AR); McKusick-Kaufman syndrome (AR) | AR | CTI | Yes |
| *MMACHC* | Methylmalonic aciduria and homocystinuria, cblC type | AR | CR | Yes |
| *MNX1* | Currarino Syndrome | AD | CS | Yes |
| *MOCOS* | Xanthinuria, Type II | AR | T | Yes |
| *MUT* | Methylmalonic Aciduria, Type mut0 (AR) | AR | G; T | Yes |
| *MVK* | Mevalonate Kinase Deficiency | AR | G | Yes |
| *MYCN* | Feingold Syndrome 1 (AD) | AD | CS | No |
| *MYH9* | MYH9-Related Disorders (AD) | AD | G | Yes |
| *MYO1E* | Focal Segmental Glomerulosclerosis, 6 | AR | G | Yes |
| *NEK8* | Nephronophthisis 9; Renal-Hepatic-Pancreatic Dysplasia | AR | CTI | Yes |
| *NEUROD1* | Maturity-Onset Diabetes of the Young 6 | AD | G | Yes |
| *NF1* | Neurofibromatosis, Type 1 | AD | G; T | Yes |
| *NLRP3* | Cryopyrin-Associated Periodic Syndromes (CAPS) (AD) | AD | G | No |
| *NOTCH2* | Acroosteolysis Dominant Type; Alagille syndrome 2; Hajdu- Cheney Syndrome | AD | CS; CTI | Yes |
| *NPHP1* | Nephronophthisis 1, Juvenile; Senior-Loken Syndrome; Joubert Syndrome 4 | AR | CTI | Yes |
| *NPHP3* | NPHP3-related conditions | AR | CTI | Yes |
| *NPHP4* | Nephronophthisis 4; Senior-Loken syndrome 4 | AR | CTI | Yes |
| *NPHS1* | Congenital Nephrotic Syndrome, Finnish Type | AR | G | Yes |
| *NPHS2* | Congenital nephrotic syndrome, type 2 | AR | G | Yes |
| *NR0B1* | Congenital Adrenal Hypoplasia with Hypogonadotropic Hypogonadism | XL | T | Yes |
| *NR3C1* | Glucocorticoid Resistance, Generalized | AD | T | Yes |
| *NR3C2* | Pseudohypoaldosteronism Type I, Autosomal Dominant Hypertension, Early-Onset | AD | T | Yes |
| *NSD1* | Sotos Syndrome | AD | CS | Yes |
| *NSDHL* | CHILD syndrome (XLD); CK syndrome (XLR) | XL | CS | Yes |
| *OCRL* | Dent disease 2; Lowe Syndrome | XL | T | Yes |
| *OFD1* | OFD1-Related Conditions (XL) | XL | CTI | Yes |
| *OPLAH* | 5-Oxoprolinase Deficiency | AR | T | Yes |
| *PAX2* | PAX2-Related Disorders (AD) | AD | CS; G | Yes |
| *PBX1* | Congenital Anomalies of the Kidney and Urinary Tract syndrome with or without Hearing Loss, Abnormal Ears, or Developmental Delay (CAKUTHED) | AD | CS | Yes |
| *PCBD1* | BH4-Deficient Hyperphenylalaninemia D (AR); Juvenile-Onset Diabetes Mellitus (AR) | AR | G; T | Yes |
| *PDSS1* | Coenzyme Q10 Deficiency, Primary, 2 | AR | G | Yes |
| *PDSS2* | Coenzyme Q10 Deficiency, Primary, 3 | AR | G | Yes |
| *PDX1* | PDX1-familial monogenic diabetes, Pancreatic agenesis 1 | AD;AR | G | Yes |
| *PET100* | Mitochondrial Complex 4 deficiency | AR | T | Yes |
| *PGK1* | Phosphoglycerate kinase 1 deficiency | XL | T | No |
| *PHEX* | Hypophosphatemic Rickets | XL | T | Yes |
| *PKD1* | Polycystic Kidney Disease 1 | AD | CTI | Yes |
| *PKD2* | Polycystic Kidney Disease 2 | AD | CTI | Yes |
| *PKHD1* | Autosomal Recessive Polycystic Kidney Disease | AR | CTI | Yes |
| *PLCE1* | Nephrotic Syndrome, Type 3 | AR | G | Yes |
| *PLG* | Congenital Plasminogen Deficiency (AR) | AR | CR | Yes |
| *PMM2* | Congenital Disorder of Glycosylation, Type 1A; Hyperinsulinemic Hypoglycemia and Polycystic Kidney Disease | AR | G | Yes |
| *PPP3CA* | Arthrogryposis, Cleft Palate, Craniosynostosis, and Impaired Intellectual Development | AD | T | No |
| *PRKCSH* | Polycystic Liver Disease 1 | AD | CTI | Yes |
| *PRODH* | Hyperprolinemia, Type 1 | AR | CS; G; T | Yes |
| *PROKR2* | Hypogonadotropic hypogonadism 3 with or without anosmia | AD;AR | CS | Yes |
| *PRPS1* | Phosphoribosylpyrophosphate synthetase superactivity | XL | T | No |
| *PTH1R* | Metaphyseal Chondrodysplasia, Murk Jansen Type (AD); Failure of tooth eruption, primary (AD); Chondrodysplasia, Blomstrand type (AR); Eiken syndrome (AR) | AD;AR | CTI | Yes |
| *PTPN11* | Noonan Syndrome (AD); Noonan Syndrome with Multiple Lentigines (AD) | AD | CS | Yes |
| *PTPRO* | Nephrotic Syndrome, Type 6 | AR | G | Yes |
| *RAD51C* | Fanconi Anemia, Group O | AR | CS | Yes |
| *REN* | Autosomal Dominant Tubulointerstitial Kidney Disease (ADTKD-REN) (AD); Renal Tubular Dysgenesis (REN-dRTD) (AR) | AD;AR | CTI | Yes |
| *RET* | Multiple endocrine neoplasia 2A/2B; Familial medullary thyroid carcinoma | AD | CS | Yes |
| *RMND1* | Combined Oxidative Phosphorylation Deficiency 11 | AR | CS | Yes |
| *ROBO2* | Congenital Anomalies of the Kidney and Urinary Tract | AD | CS | No |
| *ROR2* | Brachydactyly, type B1 (AD), Robinow Syndrome (AR) | AR | T | Yes |
| *RPGRIP1L* | Ciliopathies, RPGRIP1L-Related | AR | CTI | Yes |
| *RPL11* | Diamond-Blackfan Anemia 7 | AD | CS | Yes |
| *RPL35A* | Diamond-Blackfan Anemia 5 | AD | CS | No |
| *RPL5* | Diamond-Blackfan Anemia 6 | AD | CS | No |
| *RPS10* | Diamond-Blackfan Anemia 9 | AD | CS | No |
| *RPS17* | Diamond-Blackfan Anemia 4 | AD | CS | No |
| *RPS19* | Diamond-Blackfan Anemia 1 | AD | CS | No |
| *RPS24* | Diamond-Blackfan Anemia 3 | AD | CS | No |
| *RPS26* | Diamond-Blackfan Anemia 10 | AD | CS | No |
| *RPS7* | Diamond-Blackfan Anemia 8 | AD | CS | No |
| *RRM2B* | RRM2B Mitochondrial DNA Maintenance Defects (RRM2B-MDMDs) (AD/AR) | AD;AR | T | Yes |
| *SALL1* | Townes-Brocks syndrome 1 | AD | CS | Yes |
| *SALL4* | SALL4-Related Disorders (AD) | AD | CS | No |
| *SARS2* | Hyperuricemia, Pulmonary Hypertension, Renal Failure, And Alkalosis Syndrome (HUPRA Syndrome) (AR) | AR | T | Yes |
| *SCARB2* | Action Myoclonus-Renal Failure (AMRF) Syndrome | AR | G | Yes |
| *SCN4A* | SCN4A-Related Disorders | AD;AR | T | Yes |
| *SCNN1A* | Pseudohypoaldosteronism, Type 1 | AR | T | Yes |
| *SCNN1B* | Liddle Syndrome 1 (AD); Pseudohypoaldosteronism, Type 1 (AR) | AD;AR | T | Yes |
| *SCNN1G* | Liddle Syndrome 2 (AD); Pseudohypoaldosteronism, type 1 (AR) | AD;AR | T | Yes |
| *SCO1* | Mitochondrial Complex 4 deficiency | AR | T | Yes |
| *SDCCAG8* | Senior Loken Syndrome, Type 7; Bardet-Biedl Syndrome 16 | AR | CTI | Yes |
| *SEC63* | Polycystic Liver Disease 2 | AD | T | Yes |
| *SEMA3E* | CHARGE syndrome | UK | CS | No |
| *SI* | Sucrase-Isomaltase Deficiency | AR | T | Yes |
| *SIX1* | Branchiootorenal Spectrum Disorders (AD) | AD | CS | No |
| *SIX5* | Branchiootorenal Spectrum Disorders (AD) | AD | CS | No |
| *SLC12A1* | Bartter syndrome | AR | T | Yes |
| *SLC12A3* | Gitelman syndrome | AR | T | Yes |
| *SLC16A12* | Juvenile cataract with microcornea | AD | T | No |
| *SLC22A12* | Hypouricemia, Renal 1 | AR | T | Yes |
| *SLC26A1* | Nephrolithiasis, Calcium Oxalate | AR | T | No |
| *SLC2A2* | Fanconi-Bickel Syndrome | AR | T | Yes |
| *SLC2A9* | Renal Hypouricemia 2 | AD;AR | T | Yes |
| *SLC34A1* | Hypophosphatemic Nephrolithiasis/Osteoporosis 1 (AD) & Infantile Hypercalcemia 2 (AR) | AD;AR | T | Yes |
| *SLC34A3* | Hypophosphatemic Rickets with Hypercalciuria | AR | T | Yes |
| *SLC37A4* | Glycogen Storage Disease, Type 1B/1C (AR) | AR | G; T | Yes |
| *SLC3A1* | Cystinuria | AD;AR | T | Yes |
| *SLC4A1* | SLC4A1-associated Distal Renal Tubular Acidosis (AD/AR); Ovalocytosis, SA Type (AD); Cryohydrocytosis (AD); Spherocytosis, Type 4 (AD) | AD;AR | T | Yes |
| *SLC4A4* | Renal Tubular Acidosis, Proximal, with Ocular Abnormalities | AR | T | Yes |
| *SLC5A1* | Glucose-Galactose Malabsorption | AR | T | Yes |
| *SLC5A2* | Renal Glucosuria | AD;AR | T | Yes |
| *SLC6A19* | Hyperglycinuria (AD); Hartnup Disorder (AR); Iminoglycinuria (Complex) | AD;AR | T | Yes |
| *SLC7A7* | Lysinuric protein intolerance | AR | T | Yes |
| *SLC7A9* | Cystinuria | AD;AR | T | Yes |
| *SLX4* | Fanconi Anemia, Group P | AR | CS | Yes |
| *SMAD9* | Pulmonary Hypertension, Primary 2 | AD | T | Yes |
| *SMARCAL1* | Schimke immunoosseous dysplasia | AR | G | Yes |
| *SMC1A* | Cornelia de Lange Syndrome (XL); Developmental and Epileptic Encephalopathy 85, with or without Midline Brain Defects (DEE85) | XL | CS | Yes |
| *SOX17* | Vesicoureteral Reflux 3 | AD | CS | Yes |
| *SOX18* | SOX18-Related Disorders (AD/AR) | AD;AR | G; ? | No |
| *SRCAP* | Floating-Harbor Syndrome | AD | CS; T | Yes |
| *STAR* | Lipoid Adrenal Hyperplasia | AR | CS | Yes |
| *STX16* | Pseudohypoaldosteronism | AD | T | No |
| *TACO1* | Mitochondrial Complex 4 deficiency | AR | T | Yes |
| *TFAP2A* | Branchiooculofacial Syndrome | AD | CS | No |
| *THBD* | Atypical Hemolytic Uremic Syndrome | AD | CR | Yes |
| *TMEM67* | TMEM67-related conditions | AR | CTI | Yes |
| *TNS2* | Nephrotic Syndrome | UK | G | No |
| *TP53RK* | Galloway-Mowat Syndrome | AR | G | No |
| *TP63* | P63-Related Conditions | AD | CS | Yes |
| *TRIM32* | Limb-girdle muscular dystrophy, type 2H (AR) | AR | CTI | Yes |
| *TRPC6* | Focal Segmental Glomerulosclerosis 2 | AD | G | Yes |
| *TRPM6* | Hypomagnesemia 1, Intestinal | AR | T | Yes |
| *TSC1* | Tuberous sclerosis | AD | CTI | Yes |
| *TSC2* | Tuberous sclerosis | AD | CTI | Yes |
| *TTC21B* | Nephronophthisis 12; Short-rib thoracic dysplasia 4 with or without polydactyly | AR | CTI; G | Yes |
| *TTC8* | Bardet-Biedl Syndrome 8 | AR | CTI | Yes |
| *TXNL4A* | Burn-McKeown syndrome | AR | CS | Yes |
| *UMOD* | Autosomal Dominant Tubulointerstitial Kidney Disease (ADTKD-UMOD) (AD) | AD | CTI | Yes |
| *UPK3A* | Renal Hypodysplasia | UK | CS | No |
| *UQCC2* | Mitochondrial Complex 3 Deficiency, Nuclear Type 7 | AR | T | Yes |
| *VDR* | Vitamin D-dependent Rickets, Type 2A | AR | T | Yes |
| *VHL* | Von Hippel-Lindau Syndrome (AD); Familial Erythrocytosis 2 (AR) | AD | CTI | Yes |
| *WAS* | Wiskott-Aldrich Syndrome (XL); X-Linked Thrombocytopenia (XL); X-Linked Severe Congenital Neutropenia (XL) | XL | G | No |
| *WDPCP* | Bardet-Biedl Syndrome 15 | AR | CTI | Yes |
| *WDR19* | WDR19-related conditions (see note below) | AR | CTI | Yes |
| *WDR72* | Amelogenesis Imperfecta with or without Distal Renal Tubular Acidosis - WDR72 | AR | T | Yes |
| *WDR73* | Galloway-Mowat Syndrome | AR | G | Yes |
| *WFS1* | Wolfram Syndrome Spectrum Disorder (AD/AR) | AD;AR | CS; T | Yes |
| *WNK1* | Pseudohypoaldosteronism, Type 2C (AD); Autosomal recessive Neuropathy, hereditary sensory and autonomic, type II (AR) | AD;AR | T | Yes |
| *WNK4* | Pseudohypoaldosteronism, Type 2B | AD | T | Yes |
| *WNT4* | Mullerian Aplasia and Hyperandrogenism | AD | CS | No |
| *WNT5A* | Robinow Syndrome 1 | AD | CS | No |
| *WT1* | WT1-Related Disorders | AD | CS; G | Yes |
| *XDH* | Xanthinuria, Type I | AR | T | Yes |
| *XPNPEP3* | Nephronophthisis-Like Nephropathy 1 | AR | CTI | Yes |
| *XRCC4* | Short Stature, Microcephaly, and Endocrine Dysfunction | AR | CS | Yes |

Disease Category Abbreviation

*CTI	Cystic & Tubulointerstitial Disorders*

*G	Glomerular Disorders*

*T	Tubulopathy and Tubular Disorders*

*CS	CAKUT & Structural Disorders*

*CR	Complement-related Kidney Disorders*

**Supplemental Table 2: Comparative analysis of self-reported or designated race/ethnicity and genetic clusters derived from principal component analysis.**

| **Self-reported race/ethnicity** | **No. of Unique Samples** | | **Genetic Cluster** |  |  |  |  |  |
| --- | --- | --- | --- | --- | --- | --- | --- | --- |
|  |  | **Admixed American** | | **African** | **Central-South Asian** | **East Asian** | **European** | **Unclustered Samples*** |
| African American | 2893 | 15 | | 2718 | 2 | 0 | 26 | 132 |
| Caucasian | 5563 | 138 | | 51 | 9 | 20 | 5110 | 235 |
| East Asian | 213 | 0 | | 1 | 12 | 192 | 2 | 6 |
| Hispanic | 1912 | 1540 | | 115 | 0 | 17 | 179 | 61 |
| Mediterranean | 70 | 0 | | 1 | 7 | 1 | 58 | 3 |
| Multiples | 241 | 31 | | 53 | 8 | 23 | 116 | 10 |
| Other | 445 | 41 | | 54 | 49 | 107 | 180 | 14 |
| South-East Asian | 302 | 1 | | 0 | 135 | 147 | 8 | 11 |
| Unknown | 3542 | 547 | | 685 | 146 | 262 | 1779 | 123 |

*No. of Samples excluded during PCA filtering steps due to missing the minimum number of single-nucleotide variants after quality control.

**Supplemental Table 3: Distribution of global ancestry proportions across five continental populations in genetic clusters from principal component analysis.** Numbers displayed are median proportions and interquartile ranges.

| **Genetic Cluster** | **N** | **Global Ancestry Proportions** | | | | |
| --- | --- | --- | --- | --- | --- | --- |
|  |  | **African** | **European** | **Admixed_American** | **East Asian** | **Central-South Asian** |
| Admixed American | 2313 | 0.05 [0.02-0.09] | 0.44 [0.34-0.54] | 0.37 [0.26-0.48] | 0.06 [0.01-0.11] | 0 [0-0.08] |
| African | 3678 | 0.7 [0.63-0.76] | 0.22 [0.15-0.29] | 0.01 [0-0.06] | 0.01 [0-0.05] | 0 [0-0.04] |
| Central-South Asian | 368 | 0 [0-0.02] | 0.33 [0.26-0.43] | 0.03 [0-0.08] | 0.3 [0.22-0.36] | 0.29 [0.24-0.34] |
| East Asian | 769 | 0 [0-0] | 0.04 [0-0.09] | 0.06 [0-0.13] | 0.87 [0.78-0.93] | 0 [0-0] |
| European | 7458 | 0 [0-0.02] | 0.82 [0.75-0.88] | 0.01 [0-0.06] | 0.01 [0-0.07] | 0.1 [0.02-0.15] |

**Supplemental Table 4: Global ancestry proportions in five continental populations based on self-reported or designated race/ethnicity.** Numbers displayed are median proportions and interquartile ranges.

| **Self-reported race /ethnicity** | **N** | **Genetic cluster** | | | | |
| --- | --- | --- | --- | --- | --- | --- |
|  |  | **African** | **European** | **Admixed American** | **East Asian** | **Central-South Asian** |
| African American | 2761 | 0.71 [0.64-0.77] | 0.21 [0.15-0.28] | 0.02 [0-0.05] | 0.01 [0-0.05] | 0 [0-0.04] |
| Caucasian | 5328 | 0 [0-0.01] | 0.83 [0.76-0.89] | 0.01 [0-0.06] | 0.01 [0-0.07] | 0.09 [0.01-0.14] |
| East Asian | 207 | 0 [0-0] | 0.01 [0-0.06] | 0.06 [0.01-0.12] | 0.89 [0.81-0.94] | 0 [0-0] |
| Hispanic | 1851 | 0.06 [0.02-0.12] | 0.45 [0.34-0.55] | 0.35 [0.19-0.47] | 0.05 [0-0.11] | 0 [0-0.08] |
| Mediterranean | 67 | 0.02 [0-0.06] | 0.68 [0.59-0.76] | 0.02 [0-0.08] | 0.03 [0-0.1] | 0.15 [0.09-0.23] |
| Multiples | 231 | 0.03 [0.01-0.22] | 0.58 [0.32-0.77] | 0.03 [0-0.13] | 0.05 [0-0.11] | 0.05 [0-0.13] |
| Other | 431 | 0.01 [0-0.06] | 0.46 [0.14-0.71] | 0.04 [0-0.11] | 0.11 [0.02-0.44] | 0.05 [0-0.18] |
| South-East Asian | 291 | 0 [0-0.01] | 0.2 [0.04-0.35] | 0.04 [0-0.11] | 0.51 [0.3-0.87] | 0.05 [0-0.28] |
| Unknown | 3419 | 0.01 [0-0.11] | 0.65 [0.28-0.83] | 0.03 [0-0.12] | 0.03 [0-0.1] | 0.04 [0-0.13] |

**Supplemental Table 5: Frequency of molecular diagnosed kidney disease across the 343 genes studied, stratified by APOL1 Genotype.**

| **Gene** | **Inheritance** | **Total** | **Percentage (%)** | **High Risk APOL1 Genotype** | | **Single Risk Allele Carriers** | | **G0/G0** |  |
| --- | --- | --- | --- | --- | --- | --- | --- | --- | --- |
|  |  |  |  | **N** | **%** | **N** | **%** | **N** | **%** |
| *PKD1* | AD | 769 | 5.07 | 18 | 1.74 | 61 | 3.62 | 690 | 5.54 |
| *COL4A4* | AD;AR | 362 | 2.38 | 23 | 2.22 | 35 | 2.07 | 304 | 2.44 |
| *COL4A3* | AD;AR | 282 | 1.86 | 6 | 0.58 | 13 | 0.77 | 263 | 2.11 |
| *COL4A5* | XL | 268 | 1.77 | 9 | 0.87 | 17 | 1.01 | 242 | 1.94 |
| *PKD2* | AD | 268 | 1.77 | 3 | 0.29 | 26 | 1.54 | 239 | 1.92 |
| *HNF1B* | AD | 133 | 0.88 | 5 | 0.48 | 10 | 0.59 | 118 | 0.95 |
| *SLC12A3* | AR | 79 | 0.52 | 0 | 0 | 5 | 0.3 | 74 | 0.59 |
| *UMOD* | AD | 77 | 0.51 | 0 | 0 | 4 | 0.24 | 73 | 0.59 |
| *GLA* | XL | 50 | 0.33 | 0 | 0 | 5 | 0.3 | 45 | 0.36 |
| *SLC7A9* | AD;AR | 46 | 0.3 | 0 | 0 | 2 | 0.12 | 44 | 0.35 |
| *NPHP1* | AR | 36 | 0.24 | 0 | 0 | 1 | 0.06 | 35 | 0.28 |
| *CLCN5* | XL | 35 | 0.23 | 0 | 0 | 4 | 0.24 | 31 | 0.25 |
| *ABCC8* | AD;AR | 31 | 0.2 | 1 | 0.1 | 2 | 0.12 | 28 | 0.22 |
| *CFI* | AD;AR | 29 | 0.19 | 0 | 0 | 4 | 0.24 | 25 | 0.2 |
| *COL4A1* | AD | 29 | 0.19 | 2 | 0.19 | 0 | 0 | 27 | 0.22 |
| *HBB* | AD;AR | 28 | 0.18 | 8 | 0.77 | 4 | 0.24 | 16 | 0.13 |
| *PAX2* | AD | 28 | 0.18 | 0 | 0 | 3 | 0.18 | 25 | 0.2 |
| *NPHS2* | AR | 26 | 0.17 | 0 | 0 | 0 | 0 | 26 | 0.21 |
| *TSC2* | AD | 22 | 0.14 | 0 | 0 | 0 | 0 | 22 | 0.18 |
| *CASR* | AD;AR | 19 | 0.13 | 3 | 0.29 | 1 | 0.06 | 15 | 0.12 |
| *SLC2A9* | AD;AR | 19 | 0.13 | 0 | 0 | 2 | 0.12 | 17 | 0.14 |
| *SLC3A1* | AD;AR | 19 | 0.13 | 1 | 0.1 | 0 | 0 | 18 | 0.14 |
| *INF2* | AD | 18 | 0.12 | 0 | 0 | 0 | 0 | 18 | 0.14 |
| *CFH* | AD;AR | 17 | 0.11 | 1 | 0.1 | 1 | 0.06 | 15 | 0.12 |
| *GANAB* | AD | 16 | 0.11 | 0 | 0 | 0 | 0 | 16 | 0.13 |
| *HNF1A* | AD | 17 | 0.11 | 0 | 0 | 0 | 0 | 17 | 0.14 |
| *PRKCSH* | AD | 15 | 0.1 | 1 | 0.1 | 1 | 0.06 | 13 | 0.1 |
| *SLC4A1* | AD;AR | 15 | 0.1 | 0 | 0 | 2 | 0.12 | 13 | 0.1 |
| *TRPC6* | AD | 15 | 0.1 | 1 | 0.1 | 1 | 0.06 | 13 | 0.1 |
| *PKHD1* | AR | 14 | 0.09 | 0 | 0 | 1 | 0.06 | 13 | 0.1 |
| *CLCNKB* | AR | 12 | 0.08 | 1 | 0.1 | 0 | 0 | 11 | 0.09 |
| *FLCN* | AD | 12 | 0.08 | 1 | 0.1 | 1 | 0.06 | 10 | 0.08 |
| *WT1* | AD | 12 | 0.08 | 1 | 0.1 | 1 | 0.06 | 10 | 0.08 |
| *ALG9* | AD;AR | 10 | 0.07 | 0 | 0 | 0 | 0 | 10 | 0.08 |
| *ALPL* | AD;AR | 10 | 0.07 | 1 | 0.1 | 0 | 0 | 9 | 0.07 |
| *NR3C2* | AD | 11 | 0.07 | 0 | 0 | 1 | 0.06 | 10 | 0.08 |
| *OFD1* | XL | 10 | 0.07 | 0 | 0 | 1 | 0.06 | 9 | 0.07 |
| *PROKR2* | AD;AR | 10 | 0.07 | 1 | 0.1 | 1 | 0.06 | 8 | 0.06 |
| *SEC63* | AD | 10 | 0.07 | 1 | 0.1 | 0 | 0 | 9 | 0.07 |
| *SMAD9* | AD | 11 | 0.07 | 2 | 0.19 | 2 | 0.12 | 7 | 0.06 |
| *WFS1* | AD;AR | 10 | 0.07 | 0 | 0 | 2 | 0.12 | 8 | 0.06 |
| *AVPR2* | XL | 9 | 0.06 | 0 | 0 | 1 | 0.06 | 8 | 0.06 |
| *CEL* | AD | 9 | 0.06 | 1 | 0.1 | 2 | 0.12 | 6 | 0.05 |
| *CUBN* | AR | 9 | 0.06 | 0 | 0 | 1 | 0.06 | 8 | 0.06 |
| *GATA3* | AD | 9 | 0.06 | 0 | 0 | 1 | 0.06 | 8 | 0.06 |
| *RET* | AD | 9 | 0.06 | 0 | 0 | 1 | 0.06 | 8 | 0.06 |
| *SCNN1B* | AD;AR | 9 | 0.06 | 0 | 0 | 5 | 0.3 | 4 | 0.03 |
| *CD2AP* | AD;AR | 7 | 0.05 | 1 | 0.1 | 0 | 0 | 6 | 0.05 |
| *CTNS* | AR | 7 | 0.05 | 0 | 0 | 0 | 0 | 7 | 0.06 |
| *FGA* | AD;AR | 8 | 0.05 | 0 | 0 | 0 | 0 | 8 | 0.06 |
| *HNF4A* | AD | 7 | 0.05 | 0 | 0 | 1 | 0.06 | 6 | 0.05 |
| *LMX1B* | AD | 7 | 0.05 | 0 | 0 | 1 | 0.06 | 6 | 0.05 |
| *NF1* | AD | 7 | 0.05 | 1 | 0.1 | 1 | 0.06 | 5 | 0.04 |
| *OCRL* | XL | 8 | 0.05 | 0 | 0 | 4 | 0.24 | 4 | 0.03 |
| *PBX1* | AD | 7 | 0.05 | 0 | 0 | 0 | 0 | 7 | 0.06 |
| *SALL1* | AD | 8 | 0.05 | 0 | 0 | 0 | 0 | 8 | 0.06 |
| *SLC5A2* | AD;AR | 7 | 0.05 | 1 | 0.1 | 0 | 0 | 6 | 0.05 |
| *CYP24A1* | AR | 6 | 0.04 | 0 | 0 | 0 | 0 | 6 | 0.05 |
| *JAG1* | AD | 6 | 0.04 | 0 | 0 | 0 | 0 | 6 | 0.05 |
| *NOTCH2* | AD | 6 | 0.04 | 0 | 0 | 4 | 0.24 | 2 | 0.02 |
| *NSD1* | AD | 6 | 0.04 | 0 | 0 | 0 | 0 | 6 | 0.05 |
| *PHEX* | XL | 6 | 0.04 | 0 | 0 | 2 | 0.12 | 4 | 0.03 |
| *AGXT* | AR | 5 | 0.03 | 0 | 0 | 0 | 0 | 5 | 0.04 |
| *ANOS1* | XL | 4 | 0.03 | 2 | 0.19 | 0 | 0 | 2 | 0.02 |
| *APOA1* | AD;AR | 4 | 0.03 | 0 | 0 | 0 | 0 | 4 | 0.03 |
| *APRT* | AR | 4 | 0.03 | 0 | 0 | 1 | 0.06 | 3 | 0.02 |
| *ATP6V0A4* | AR | 5 | 0.03 | 0 | 0 | 0 | 0 | 5 | 0.04 |
| *ATP7B* | AR | 4 | 0.03 | 0 | 0 | 0 | 0 | 4 | 0.03 |
| *C3* | AD;AR | 4 | 0.03 | 0 | 0 | 0 | 0 | 4 | 0.03 |
| *EYA1* | AD | 5 | 0.03 | 1 | 0.1 | 1 | 0.06 | 3 | 0.02 |
| *GCK* | AD;AR | 4 | 0.03 | 0 | 0 | 0 | 0 | 4 | 0.03 |
| *KLHL3* | AD;AR | 5 | 0.03 | 0 | 0 | 0 | 0 | 5 | 0.04 |
| *LMNA* | AD;AR | 5 | 0.03 | 0 | 0 | 0 | 0 | 5 | 0.04 |
| *MEFV* | AR | 4 | 0.03 | 0 | 0 | 0 | 0 | 4 | 0.03 |
| *NPHP4* | AR | 4 | 0.03 | 0 | 0 | 0 | 0 | 4 | 0.03 |
| *PTPN11* | AD | 4 | 0.03 | 1 | 0.1 | 0 | 0 | 3 | 0.02 |
| *SLC12A1* | AR | 4 | 0.03 | 0 | 0 | 0 | 0 | 4 | 0.03 |
| *SLC34A1* | AD;AR | 4 | 0.03 | 0 | 0 | 0 | 0 | 4 | 0.03 |
| *VHL* | AD | 4 | 0.03 | 1 | 0.1 | 0 | 0 | 3 | 0.02 |
| *CDC73* | AD | 3 | 0.02 | 0 | 0 | 0 | 0 | 3 | 0.02 |
| *CNNM2* | AD;AR | 3 | 0.02 | 0 | 0 | 0 | 0 | 3 | 0.02 |
| *FAN1* | AR | 3 | 0.02 | 0 | 0 | 0 | 0 | 3 | 0.02 |
| *FGFR1* | AD | 3 | 0.02 | 1 | 0.1 | 0 | 0 | 2 | 0.02 |
| *FOXC1* | AD | 3 | 0.02 | 0 | 0 | 0 | 0 | 3 | 0.02 |
| *GNAS* | AD | 3 | 0.02 | 1 | 0.1 | 0 | 0 | 2 | 0.02 |
| *GRHPR* | AR | 3 | 0.02 | 0 | 0 | 0 | 0 | 3 | 0.02 |
| *INS* | AD;AR | 3 | 0.02 | 0 | 0 | 0 | 0 | 3 | 0.02 |
| *MYH9* | AD | 3 | 0.02 | 0 | 0 | 0 | 0 | 3 | 0.02 |
| *NPHS1* | AR | 3 | 0.02 | 0 | 0 | 0 | 0 | 3 | 0.02 |
| *NR3C1* | AD | 3 | 0.02 | 0 | 0 | 0 | 0 | 3 | 0.02 |
| *PDX1* | AD;AR | 3 | 0.02 | 0 | 0 | 0 | 0 | 3 | 0.02 |
| *REN* | AD;AR | 3 | 0.02 | 0 | 0 | 0 | 0 | 3 | 0.02 |
| *SCN4A* | AD;AR | 3 | 0.02 | 0 | 0 | 1 | 0.06 | 2 | 0.02 |
| *TSC1* | AD | 3 | 0.02 | 0 | 0 | 0 | 0 | 3 | 0.02 |
| *ABCC6* | AR | 1 | 0.01 | 0 | 0 | 0 | 0 | 1 | 0.01 |
| *ACTN4* | AD | 1 | 0.01 | 0 | 0 | 0 | 0 | 1 | 0.01 |
| *ADA2* | AR | 1 | 0.01 | 0 | 0 | 0 | 0 | 1 | 0.01 |
| *AQP2* | AD;AR | 2 | 0.01 | 0 | 0 | 0 | 0 | 2 | 0.02 |
| *ATP6V1B1* | AR | 2 | 0.01 | 0 | 0 | 0 | 0 | 2 | 0.02 |
| *AVP* | AD | 1 | 0.01 | 0 | 0 | 1 | 0.06 | 0 | 0 |
| *BBS1* | AR | 1 | 0.01 | 0 | 0 | 0 | 0 | 1 | 0.01 |
| *BBS10* | AR | 1 | 0.01 | 0 | 0 | 0 | 0 | 1 | 0.01 |
| *BBS2* | AR | 1 | 0.01 | 0 | 0 | 0 | 0 | 1 | 0.01 |
| *BBS9* | AR | 2 | 0.01 | 0 | 0 | 0 | 0 | 2 | 0.02 |
| *BMPR2* | AD | 2 | 0.01 | 0 | 0 | 0 | 0 | 2 | 0.02 |
| *BSCL2* | AD;AR | 1 | 0.01 | 0 | 0 | 0 | 0 | 1 | 0.01 |
| *CA2* | AR | 1 | 0.01 | 0 | 0 | 0 | 0 | 1 | 0.01 |
| *CACNA1S* | AD;AR | 1 | 0.01 | 0 | 0 | 0 | 0 | 1 | 0.01 |
| *CAV1* | AD;AR | 1 | 0.01 | 0 | 0 | 0 | 0 | 1 | 0.01 |
| *CEP290* | AR | 2 | 0.01 | 0 | 0 | 1 | 0.06 | 1 | 0.01 |
| *CHD7* | AD | 1 | 0.01 | 0 | 0 | 0 | 0 | 1 | 0.01 |
| *CLCN2* | AD;AR | 1 | 0.01 | 0 | 0 | 0 | 0 | 1 | 0.01 |
| *CLDN19* | AR | 1 | 0.01 | 0 | 0 | 0 | 0 | 1 | 0.01 |
| *CPLANE1* | AR | 1 | 0.01 | 0 | 0 | 0 | 0 | 1 | 0.01 |
| *CUL3* | AD | 2 | 0.01 | 0 | 0 | 0 | 0 | 2 | 0.02 |
| *CYP11B1* | AD;AR | 1 | 0.01 | 0 | 0 | 1 | 0.06 | 0 | 0 |
| *CYP27B1* | AR | 1 | 0.01 | 0 | 0 | 0 | 0 | 1 | 0.01 |
| *FAM20A* | AR | 2 | 0.01 | 0 | 0 | 1 | 0.06 | 1 | 0.01 |
| *FANCC* | AR | 1 | 0.01 | 0 | 0 | 0 | 0 | 1 | 0.01 |
| *FGF23* | AD;AR | 1 | 0.01 | 0 | 0 | 0 | 0 | 1 | 0.01 |
| *FGFR2* | AD | 1 | 0.01 | 0 | 0 | 0 | 0 | 1 | 0.01 |
| *FN1* | AD | 2 | 0.01 | 0 | 0 | 0 | 0 | 2 | 0.02 |
| *FOXC2* | AD | 1 | 0.01 | 0 | 0 | 0 | 0 | 1 | 0.01 |
| *FOXP3* | XL | 2 | 0.01 | 0 | 0 | 1 | 0.06 | 1 | 0.01 |
| *G6PC* | AR | 2 | 0.01 | 0 | 0 | 0 | 0 | 2 | 0.02 |
| *GALNT3* | AR | 1 | 0.01 | 0 | 0 | 0 | 0 | 1 | 0.01 |
| *GLI3* | AD | 1 | 0.01 | 0 | 0 | 0 | 0 | 1 | 0.01 |
| *GPC3* | XL | 1 | 0.01 | 0 | 0 | 1 | 0.06 | 0 | 0 |
| *GSN* | AD | 1 | 0.01 | 0 | 0 | 0 | 0 | 1 | 0.01 |
| *HOGA1* | AR | 2 | 0.01 | 0 | 0 | 0 | 0 | 2 | 0.02 |
| *HOXA13* | AD | 1 | 0.01 | 0 | 0 | 0 | 0 | 1 | 0.01 |
| *HSD11B2* | AR | 1 | 0.01 | 0 | 0 | 0 | 0 | 1 | 0.01 |
| *IFT140* | AR | 2 | 0.01 | 0 | 0 | 0 | 0 | 2 | 0.02 |
| *IQCB1* | AR | 2 | 0.01 | 0 | 0 | 0 | 0 | 2 | 0.02 |
| *KANSL1* | AD | 1 | 0.01 | 0 | 0 | 0 | 0 | 1 | 0.01 |
| *KCNJ1* | AR | 2 | 0.01 | 0 | 0 | 0 | 0 | 2 | 0.02 |
| *KCNJ11* | AD;AR | 1 | 0.01 | 1 | 0.1 | 0 | 0 | 0 | 0 |
| *LRP5* | AD;AR | 1 | 0.01 | 0 | 0 | 0 | 0 | 1 | 0.01 |
| *MAFB* | AD | 1 | 0.01 | 0 | 0 | 0 | 0 | 1 | 0.01 |
| *MMACHC* | AR | 1 | 0.01 | 0 | 0 | 0 | 0 | 1 | 0.01 |
| *MNX1* | AD | 1 | 0.01 | 0 | 0 | 1 | 0.06 | 0 | 0 |
| *NEUROD1* | AD | 1 | 0.01 | 0 | 0 | 0 | 0 | 1 | 0.01 |
| *NPHP3* | AR | 1 | 0.01 | 0 | 0 | 0 | 0 | 1 | 0.01 |
| *NR0B1* | XL | 1 | 0.01 | 0 | 0 | 0 | 0 | 1 | 0.01 |
| *NSDHL* | XL | 1 | 0.01 | 0 | 0 | 0 | 0 | 1 | 0.01 |
| *PMM2* | AR | 1 | 0.01 | 0 | 0 | 0 | 0 | 1 | 0.01 |
| *PTH1R* | AD;AR | 2 | 0.01 | 0 | 0 | 0 | 0 | 2 | 0.02 |
| *RMND1* | AR | 1 | 0.01 | 0 | 0 | 0 | 0 | 1 | 0.01 |
| *RPL11* | AD | 1 | 0.01 | 0 | 0 | 0 | 0 | 1 | 0.01 |
| *RRM2B* | AD;AR | 1 | 0.01 | 0 | 0 | 0 | 0 | 1 | 0.01 |
| *SCARB2* | AR | 2 | 0.01 | 0 | 0 | 1 | 0.06 | 1 | 0.01 |
| *SCNN1A* | AR | 1 | 0.01 | 0 | 0 | 0 | 0 | 1 | 0.01 |
| *SCNN1G* | AD;AR | 1 | 0.01 | 0 | 0 | 0 | 0 | 1 | 0.01 |
| *SDCCAG8* | AR | 2 | 0.01 | 0 | 0 | 0 | 0 | 2 | 0.02 |
| *SMC1A* | XL | 1 | 0.01 | 0 | 0 | 0 | 0 | 1 | 0.01 |
| *SOX17* | AD | 1 | 0.01 | 0 | 0 | 0 | 0 | 1 | 0.01 |
| *SRCAP* | AD | 1 | 0.01 | 0 | 0 | 0 | 0 | 1 | 0.01 |
| *THBD* | AD | 1 | 0.01 | 0 | 0 | 0 | 0 | 1 | 0.01 |
| *TMEM67* | AR | 1 | 0.01 | 0 | 0 | 0 | 0 | 1 | 0.01 |
| *TP63* | AD | 1 | 0.01 | 0 | 0 | 0 | 0 | 1 | 0.01 |
| *TRPM6* | AR | 1 | 0.01 | 0 | 0 | 0 | 0 | 1 | 0.01 |
| *TTC21B* | AR | 1 | 0.01 | 0 | 0 | 0 | 0 | 1 | 0.01 |
| *TXNL4A* | AR | 1 | 0.01 | 0 | 0 | 0 | 0 | 1 | 0.01 |
| *WDR19* | AR | 1 | 0.01 | 0 | 0 | 0 | 0 | 1 | 0.01 |
| *WNK4* | AD | 1 | 0.01 | 0 | 0 | 0 | 0 | 1 | 0.01 |

**Supplemental Table 6: Detailed Characteristics of Samples with *APOL1* Risk Alleles**

| Characteristic | High Risk Genotype | Single Risk Allele Carriers |
| --- | --- | --- |
| Self-reported race & ethnicity |  |  |
| African American | 802 | 1137 |
| Caucasian | 6 | 36 |
| East Asian | 0 | 0 |
| Hispanic | 36 | 127 |
| Mediterranean | 0 | 0 |
| Multiples | 13 | 23 |
| Other | 17 | 23 |
| South-East Asian | 0 | 0 |
| Unknown | 161 | 341 |
| Genetic cluster |  |  |
| Admixed American | 27 | 141 |
| African | 944 | 1453 |
| Central-South Asian | 0 | 2 |
| East Asian | 0 | 1 |
| European | 4 | 37 |
| Ancestry Proportions |  |  |
| Admixed American | 0.02 [0-0.06] | 0.02 [0-0.07] |
| African | 0.71 [0.65-0.77] | 0.69 [0.59-0.75] |
| Central-South Asian | 0 [0-0.04] | 0 [0-0.04] |
| East Asian | 0.01 [0-0.05] | 0.01 [0-0.06] |
| European | 0.21 [0.15-0.28] | 0.23 [0.16-0.32] |

**Supplemental Table 7: Detailed Characteristics of Samples Selected by Matching Analysis**

| **Categories** | **All Patients** | **High Risk Genotype** | **Single Risk Genotype** | **G0/G0** |
| --- | --- | --- | --- | --- |
| No. of Samples | 2890 | 975 | 952 | 963 |
| Gender & Age |  |  |  |  |
| Female | 1335 (46.2) | 446 (45.7) | 433 (45.5) | 456 (47.4) |
| Male | 1555 (53.8) | 529 (54.3) | 519 (54.5) | 507 (52.6) |
| Age yr [IQR] | 46 [31-59] | 45 [32-57] | 46 [32-60] | 47 [28.5-62] |
| Mendelian Disease Status |  |  |  |  |
| Affected | 376 (13) | 92 (9.4) | 128 (13.4) | 156 (16.2) |
| Carrier | 1000 (34.6) | 354 (36.3) | 336 (35.3) | 310 (32.2) |
| Negative | 1514 (52.4) | 529 (54.3) | 488 (51.3) | 497 (51.6) |
| Self-reported race & ethnicity |  |  |  |  |
| African American | 2158 (74.7) | 749 (76.8) | 708 (74.4) | 701 (72.8) |
| Caucasian | 29 (1) | 6 (0.6) | 7 (0.7) | 16 (1.7) |
| Hispanic | 102 (3.5) | 33 (3.4) | 28 (2.9) | 41 (4.3) |
| Multiples | 38 (1.3) | 13 (1.3) | 10 (1.1) | 15 (1.6) |
| Others | 44 (1.5) | 17 (1.7) | 7 (0.7) | 20 (2.1) |
| Unknown | 519 (18) | 157 (16.1) | 192 (20.2) | 170 (17.7) |
| Genomic Ancestry Proportions |  |  |  |  |
| African | 0.71 [0.65-0.77] | 0.71 [0.65-0.77] | 0.72 [0.65-0.77] | 0.71 [0.64-0.77] |
| Admixed American | 0.02 [0-0.06] | 0.02 [0-0.06] | 0.02 [0-0.06] | 0.02 [0-0.06] |
| Central-South Asian | 0 [0-0.04] | 0 [0-0.04] | 0 [0-0.04] | 0 [0-0.04] |
| East Asian | 0.01 [0-0.05] | 0.01 [0-0.05] | 0.01 [0-0.05] | 0.01 [0-0.05] |
| European | 0.21 [0.15-0.28] | 0.21 [0.15-0.28] | 0.21 [0.14-0.28] | 0.21 [0.15-0.28] |
| Genetic Cluster |  |  |  |  |
| Admixed American | 73 (2.5) | 27 (2.8) | 26 (2.7) | 20 (2.1) |
| African | 2791 (96.6) | 944 (96.8) | 920 (96.6) | 927 (96.3) |
| Central-South Asian | 1 (0) | 0 (0) | 1 (0.1) | 0 (0) |
| European | 25 (0.9) | 4 (0.4) | 5 (0.5) | 16 (1.7) |
| Chronic kidney disease | 1675 | 547 (32.7) | 561 (33.5) | 567 (33.9) |
| End-stage kidney disease | 473 | 226 (47.8) | 133 (28.1) | 114 (24.1) |
| Unclustered samples | 225 | 68 (30.2) | 77 (34.2) | 80 (35.6) |
| Clinical diagnosis (ICD codes)& |  |  |  |  |
| Proteinuria/nephrotic syndrome | 223 | 74 (33.2) | 77 (34.5) | 72 (32.3) |
| Cystic kidney disease | 168 | 33 (19.6) | 67 (39.9) | 68 (40.5) |
| Others | 148 | 22 (14.9) | 56 (37.8) | 70 (47.3) |
| Hypertension | 107 | 27 (25.2) | 36 (33.6) | 44 (41.1) |
| Kidney transplant | 60 | 26 (43.3) | 17 (28.3) | 17 (28.3) |
| Nephritic syndrome | 59 | 32 (54.2) | 12 (20.3) | 15 (25.4) |
| Hematuria | 58 | 16 (27.6) | 23 (39.7) | 19 (32.8) |
| Kidney stone | 4 | 3 (75) | 0 (0) | 1 (25) |
